# Genome-wide association analyses reveal susceptibility variants linked to Parkinson’s disease in the South African population using inferred global and local ancestry

**DOI:** 10.1101/2025.08.01.25331910

**Authors:** Kathryn Step, Thiago Peixoto Leal, Emily Waldo, Lusanda Madula, Yolandi Swart, Carlos F. Hernández, Jonggeol Jeffrey Kim, Sara Bandres-Ciga, Global Parkinson’s Genetics Program (GP2), Ignacio F. Mata, Soraya Bardien

## Abstract

Genome-wide association studies (GWAS) have been successful in identifying over 100 loci associated with Parkinson’s disease (PD) susceptibility. However, the majority of these studies have focused on European cohorts with few including diverse ancestries. Using genotyped and imputed data from 691 South African PD cases and 826 controls, we conducted a conventional GWAS, two local ancestry GWAS (LA-GWAS) approaches (one using local ancestry as a covariate and the other separating the dosage per ancestry), and an association analysis to identify regions of homozygosity associated with PD status. Furthermore, we replicated these findings using another admixed population, a Latin American cohort (LARGE-PD). The ancestry inference suggested that the South African cohort is admixed from five populations, including African (AFR), European (EUR), Malaysian (MAL), Nama (NAMA), and South Asian (SAS), though with varying accuracy levels. The conventional GWAS successfully identified one locus (rs17098735-T) with genome-wide significance (p-value: 1.23×10^-8^; beta= 2.286; SE= 0.401). Within the local ancestry window of the top GWAS hit, among individuals carrying the variant, 86.7% had AFR ancestry, 11% NAMA ancestry, and 2.2% MAL ancestry, with no EUR or SAS ancestry observed, highlighting a potential ancestry-specific genetic risk factor. Three lead loci were replicated in the LARGE-PD cohort. LA-GWAS using the Cochran-Armitage trend test identified 35 lead SNPs above suggestive significance after multiple test correction. Tractor-based approaches identified three lead loci when analyzing all five ancestry components jointly, as well as three additional lead loci in the AFR-only component, highlighting ancestry-specific loci that may contribute to genetic risk in diverse populations. For the LA-GWAS, one independent locus was replicated in LARGE-PD. Our findings suggest ancestry specificity in PD risk and underscores the importance of including diverse populations in genetics research. The study contributes towards a global understanding of the genetic etiology underlying PD.

## Introduction

Stemming from a complex etiology that includes a strong genetic component (Trevisan *et al*., 2024), Parkinson’s disease (PD) is a neurodegenerative disorder characterized by a wide range of both motor and non-motor symptoms (Armstrong and Okun, 2020). The burden of PD in Sub-Saharan African populations is increasing rapidly within the aging populations, ranking as the 11^th^ most prevalent nervous system disorder in the region (GBD 2021 Nervous System Disorders Collaborators, 2024). By 2040, it is predicted that there will be 12 million individuals with PD globally (Dorsey *et al*., 2018; Karikari *et al*., 2018), with Africa expected to face the greatest impact (Dorsey *et al*., 2007). However, despite the rise in disease prevalence, the representation of African populations in PD genetic research remains limited.

The South African population is genetically diverse and comprises several diverse global ancestries, including the African, Asian, and European groups (Thami and Chimusa, 2019). Notably, southern Africa is home to indigenous populations, such as the Khoe-San, who possess some of the most divergent lineages with shorter linkage disequilibrium (LD) blocks (Henn *et al*., 2011; Schlebusch *et al*., 2012). Ancestral allele frequencies vary across genomes due to factors such as natural selection, genetic drift, and differing exposures to environments and pathogens (Brown *et al*., 2017; Schubert, Andaleon and Wheeler, 2020). These variations in allele frequencies can help identify population-specific disease risk variants in admixed individuals while simultaneously uncovering risk variants relevant to multiple populations (Swart *et al*., 2021).

Previous PD genome-wide association studies (GWAS) and meta-analyses have identified 90 independent risk loci in a European cohort (Nalls *et al*., 2019), 134 loci through a European meta-analysis (Global Parkinson’s Genetics Program (GP2) and Leonard, 2025), 11 in an Asian cohort (Foo *et al*., 2020), and one in a Latin American cohort (Loesch *et al*., 2021). Recent efforts to diversify PD genetic research include a multiple-ancestry meta-analysis, comprising ancestral groups previously underrepresented, which identified 78 independent loci with 12 potentially novel loci (Kim *et al*., 2024). The first PD GWAS that was focused on African and African admixed participants successfully identified a novel ancestry-specific risk variant (rs3115534-G) in the *GBA1* gene (Rizig *et al*., 2023).

Here, we aim to conduct the first GWAS for PD in a South African study cohort with computational approaches best suited for multi-way admixed populations including a GWAS incorporating local ancestry, which refers to the genetic contribution from specific ancestral populations at an individual level (Fu and Shi, 2024). The inclusion of this diverse ancestral population has the potential to uncover additional genetic risk factors on a global and local ancestral scale.

## Methods and materials

An overview of the methods is provided in **Figure 1**.

**Figure 1:**
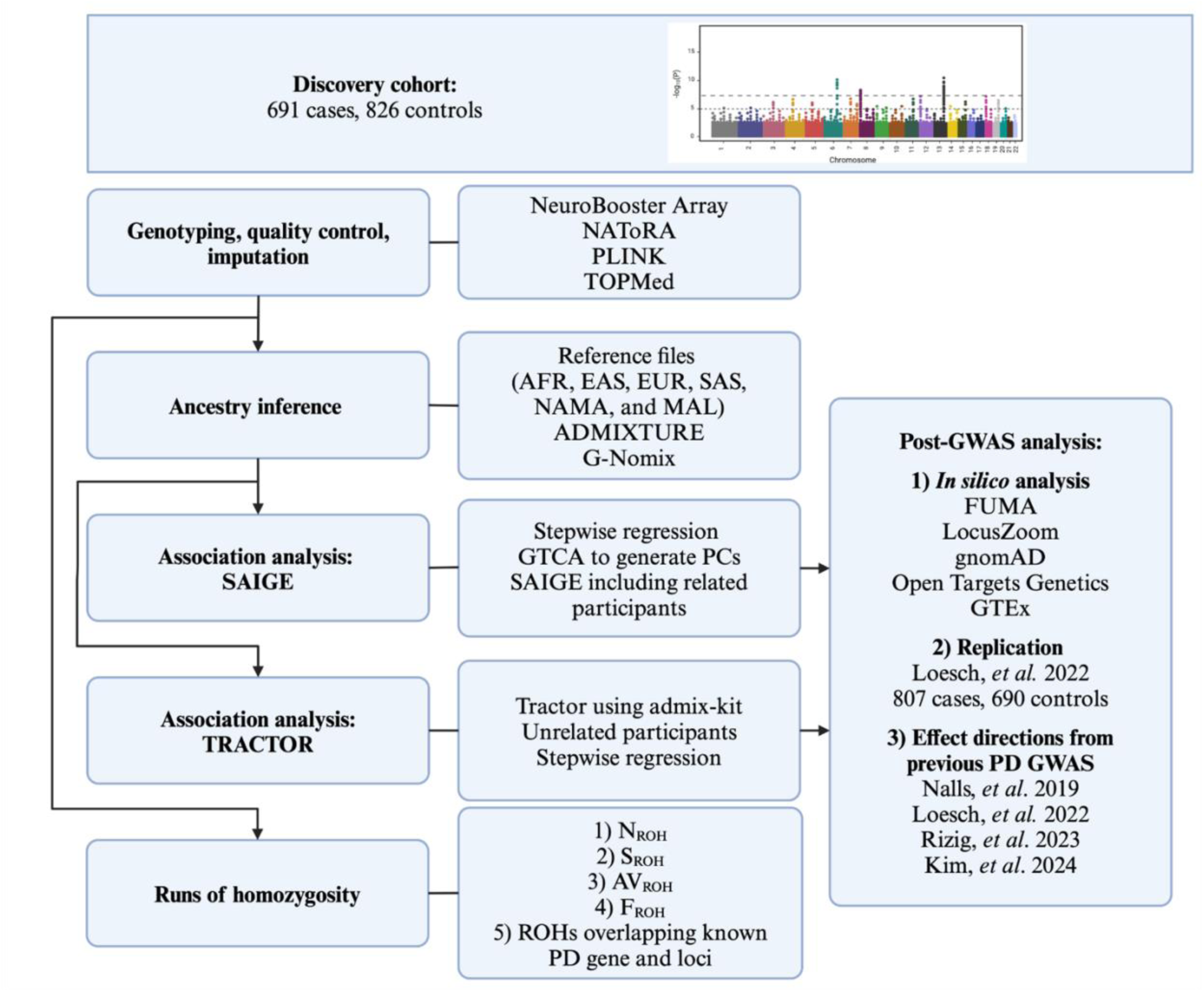
Overview of the methods followed in the present study. AFR, African; AV_ROH_, average length of ROHs; EAS, East Asian; EUR, European; F_ROH_, Inbreeding coefficient estimates based on ROHs; FUMA, Functional Mapping and Annotation; MAL, Malaysian; NAMA, Nama-Khoe; N_ROH_, number of ROHs; PCs, Principal components; PD, Parkinson’s disease; ROH, runs of homozygosity; SAIGE, Scalable and Accurate Implementation of GEneralized mixed model; SAS, South Asian; S_ROH_, sum or total length of ROHs

### Study population and ethics approval

For the discovery cohort, 691 individuals living with PD, diagnosed by a neurologist specializing in movement disorders according to the Queen’s Square Brain Bank Criteria (Hughes *et al*., 1992), were recruited from 2002 until June 2020 as part of the South African Parkinson’s Disease Study Collection (SAPDSC) (van Rensburg *et al*., 2022). A total of 826 controls were recruited from 2005 until 2018 (Health Research Ethics Committee, Stellenbosch University, 2002C/059). Participants (**Supplementary Table 1**) were submitted to the Global Parkinson’s Genetics Program (GP2) for genotyping (Global Parkinson’s Genetics Program, 2021; Step, Eltaraifee, *et al*., 2025).

For replication, we used an independent cohort from the Latin American Research consortium on the Genetics of Parkinson’s Disease (LARGE-PD) Phase 1 (**Supplementary Table 2**), as previously described (Loesch *et al*., 2021). Full details on the cohort recruitment (807 PD cases and 690 controls), genotyping with Illumina’s Multi-Ethnic Genotyping Array (MEGA), quality control, and imputation can be found in Loesch, *et al*. 2021. This cohort was selected due to its high level of admixture, similar to the South African population, though the specific ancestral contributions differ (Norris *et al*., 2018; Loesch *et al*., 2021). Both populations exhibit greater admixture compared to other global populations.

### Genotyping, quality control, and imputation

Genotyping of the SAPDSC was performed by GP2 using the NeuroBooster Array (v1.0, Illumina, San Diego, CA), which is composed of a backbone of 1,914,934 genome-wide variants and 95,273 variants associated with neurological diseases or traits (Bandres-Ciga *et al*., 2023). Initial quality control (QC) was performed using PLINK v1.9 (Purcell *et al*., 2007) and PLINK v2.0 (Chang *et al*., 2015), as previously described (Leal *et al*., 2025). The QC steps included filtering for Hardy-Weinberg equilibrium (1×10^-10^ in cases, 1×10^-6^ in controls), excess heterozygosity (>3 standard deviations), removing duplicate variants, data missingness at genotype and individual level (5%), redundant variants (A/T or C/G), and sex discrepancies (Females: F <0.5, Males: F >0.8). The QC also identified a list of related individuals using King v2.2.9 (Manichaikul *et al*., 2010) and a kinship coefficient cutoff of 0.0884 indicating second degree relation (Manichaikul *et al*., 2010). Related individuals were excluded using NAToRA (Leal *et al*., 2022) (**Supplementary Figure 1**). These individuals were excluded at the relevant steps in the downstream analysis where relatedness was not permitted, while they were retained for analyses that allowed related individuals. Overall, 1,245,359 variants and 1,461 samples passed QC (679 cases; 782 controls). The Trans-Omics for Precision Medicine (TOPMed) Imputation Server was used for imputation with panel R3 (Das *et al*., 2016), and a Rsq filter of 0.3 was applied. Imputation quality was assessed using the empirical dosage Rsq (EmpRsq) values, which offer a more accurate measure than model-based Rsq by directly comparing observed and imputed genotypes. Unlike Rsq, which estimates quality based on the imputation model, EmpRsq evaluates the correlation between actual genotyped variants and their imputed counterparts, ensuring a more reliable assessment (**Supplementary Figure 2**).

### Ancestry inference

To accurately infer the ancestral proportions of the SAPDSC, we performed ancestry inference as previously described (Leal *et al*., 2025). For this, a reference panel were created using the continental non-hunter-gatherer African (AFR; n=471), continental East Asian (EAS; n=316), continental European (EUR; n=136), and continental South Asian (SAS; n=67) samples from the 1000 Genomes Project Phase III (Byrska-Bishop *et al*., 2022; Shriner *et al*., 2023). Furthermore, we made use of whole-genome sequencing data for 84 unrelated healthy controls from the Nama (NAMA) population, an indigenous hunter-gatherer Khoe-San population of Southern Africa, representing a distinct subcontinental genetic ancestry (van Eeden *et al*., 2022; Ragsdale *et al*., 2023), as well as genotyping data for 40 unrelated, healthy Malaysian (MAL) controls, also representing a subcontinental ancestral group (**Supplementary Table 3**), from GP2’s seventh data release (Leonard *et al*., 2024). These populations were included to account for the extensive genetic diversity found in the SAPDSC. A principal component analysis (PCA) was completed to identify outliers (**Supplementary Figure 3**). The reference files were phased using the TOPMed Imputation Server (Das *et al*., 2016) to ensure phasing accuracy and increase comparability to imputed data, leveraging its large and diverse reference panel for improved haplotype resolution. Population substructure was investigated using ADMIXTURE v1.3.0 (Alexander and Lange, 2011), using AFR, EUR, EAS, NAMA, and SAS for reference populations. For this, LD pruning was performed using a sliding window of 200 SNPs, with a step size of 50 SNPs, and an r² threshold of 0.2 to remove variants in high LD, where 508,428 variants were retained to infer ancestry in a supervised manner. The global ancestry proportions were visualised using PONG software v1.5 (Behr *et al*., 2016). Local ancestry (LA) was inferred using G-Nomix v1.0 (Hilmarsson *et al*., 2021) and the data phased using the TOPMed Imputation Server (Das *et al*., 2016).

### Conventional association analysis

The conventional association was conducted using methods previously described (Leal *et al*., 2025). We performed a supervised principal component analysis (PCA) using the previously mentioned parental populations to access the principal components (PCs). The SAPDSC genotyped data was then projected onto these components. A Manhattan plot for the projected PCs (1-50) was created to assess if the PCs had genome-wide influence or if they were driven by a specific genomic region (**Supplementary Figure 4**). A stepwise regression was used to identify PCs that significantly contributed to explain variance in the phenotype while minimizing redundancy. The selected PCs capture key ancestry-related variation and potential confounding effects. The analysis included the following covariates: age, sex, PC1, PC3, PC7, PC10, PC12, PC15, PC16, PC19, PC29, PC37, PC42, PC45, PC46, PC49, PC50. A regression analysis was run with Scalable and Accurate Implementation of GEneralized mixed model (SAIGE) v1.3.1 (Zhou *et al*., 2018). SAIGE was used for the association analysis due to its ability to infer and account for relatedness using the genetic relationship matrix as a random effect, enabling the retention of more individuals in the analysis. Furthermore, using conditional analysis, SAIGE can reduce the occurrence of type 1 errors, allowing for the identification of rare-variant associations while accounting for an unbalanced number of cases and controls (Zhou *et al*., 2018). An additional analysis was run using PC1-10 including a QQ plot and genomic inflation factor (**λ**GC) (**Supplementary Figure 5**). The G-Nomix output was used to infer the LA of participants carrying the top ten GWAS hits. The variant-containing window was extracted from the imputed files and matched it to the inferred ancestry in the marginal segment probability (MSP) file. Post-GWAS *in silico* analysis was conducted using LocusZoom v0.13.0 (Pruim *et al*., 2010), Functional Mapping and Annotation (FUMA) v1.3.8 (Watanabe *et al*., 2017), gnomAD v4.1.0 (Chen *et al*., 2024), Open Targets Genetics v22.10 (Ghoussaini *et al*., 2021; Mountjoy *et al*., 2021), and the Genotype-Tissue Expression (GTEx) Portal v10 (GTEx Consortium, 2013). To validate the top GWAS hit, we performed Sanger sequencing on a subset of samples carrying the variant. Polymerase chain reaction amplification and sequencing were conducted using standard protocols (detailed in **Supplementary Methods**). Sequencing chromatograms were manually reviewed to confirm the variant’s presence (**Supplementary Figure 6**).

### Local ancestry association analysis

A LA genome-wide association analysis (LA-GWAS) was run on unrelated individuals using Admix-kit v0.1.3 (Hou *et al*., 2024) and the LA results obtained from G-Nomix v1.0 (Hilmarsson *et al*., 2021), following methods previously described (Leal *et al*., 2025). Admix-kit allows for multiple association analysis models to be run simultaneously. For this analysis we ran the following: (1) Cochran-Armitage trend test (ATT; (Cochran, 1954; Zheng and Gastwirth, 2006)) and (2) Tractor (Atkinson *et al*., 2021). ATT makes use of LA information as a covariate, ultimately increasing the power of the study by 25% (Hou *et al*., 2021). Conversely, Tractor separates the dosage information per ancestry and estimates the effect sizes contributing to trait variation, increasing the power of detecting ancestry-specific associations (Atkinson *et al*., 2021). Related individuals were removed from the imputed and MSP files according to the kinship estimates completed in QC. The imputed VCF files were filtered to retain variants with a minor allele frequency (MAF) >0.005, thereby removing monomorphic and very rare variants and converted to PLINK 2.0 binary format for downstream analysis. Both models were run through Admix-kit using the stepwise regression PCs from the GWAS with SAIGE. However, for the LA-GWAS, an analysis using PC1-10 was not performed as the cohort’s complex admixture led to model overfitting, preventing p-value calculations. The ATT output files contain the beta, standard error (SE), and p-value for each variant per chromosome. The Tractor output files contained association results for each variant per ancestry as well as the results for the combined dataset, allowing for cross-ancestry comparison. Following the LA-GWAS, the results were run through the above mentioned *in silico* approaches.

### Effect directions from previous Parkinson’s disease association studies

Beta and p-values were obtained for Nalls *et al*., 2019 and Kim *et al*., 2024 from supplementary materials, Rizig *et al*., 2023 from the Neurodegenerative Disease Knowledge Portal (Dilliott *et al*., 2024), and for Loesch *et al*., 2021 from LARGE-PD. Where necessary, LiftOver v1.0 from the University of California Santa Cruz Genome Browser was used to convert to GRCh38 (Perez *et al*., 2025). Beta and p-values were extracted per positional coordinate and all reference and alternate alleles were verified against their respective alleles. Beta values were used to compare studies, as they provide a standardized measure of effect size and direction across different datasets. To ensure consistency, beta values from the previous GWAS were inverted when the effect and other alleles were in the opposite orientation from the SAPDSC GWAS, allowing for direct comparison across datasets.

### Runs of homozygosity analysis

Runs of homozygosity (ROH) are regions of consecutive homozygous genotypes (Szpiech *et al*., 2013) which typically span more than 1.5Mb (Moreno-Grau *et al*., 2021). Previous research has shown an increased homozygosity in neurodegenerative diseases, such as PD (Simón-Sánchez *et al*., 2012) and Alzheimer’s disease (Ghani *et al*., 2015; Moreno-Grau *et al*., 2021). Given that conventional GWAS primarily detects associations at the single-variant level, ROH analysis serves as a complementary approach by identifying risk loci where homozygosity may contribute to PD susceptibility. By incorporating both GWAS and ROH-based association analysis, we aimed to capture a broader spectrum of genetic risk factors, including both common and recessively inherited variants. To investigate the association of ROH with PD status, we used PLINK v1.9 (Purcell *et al*., 2007) and the methods previously described (Step, Hernández, *et al*., 2025). The analysis included assessing four ROH parameters: (1) the sum of ROHs (S_ROH_), (2) the number of ROHs (N_ROH_), (3) the average length of ROHs (AV_ROH_), and (4) the inbreeding coefficient (F_ROH_) estimates based on the ROHs. Furthermore, we investigated ROHs which overlap known PD, pallido-pyramidal syndrome, and atypical parkinsonism genes and risk loci (**Supplementary Table 4**). For this analysis, a 1Mb window downstream and upstream from the risk loci and genes were included, encompassing the 135 lead loci identified from the standard GWAS performed on the SAPDSC. Bonferroni threshold was adjusted by dividing 0.05 by the number of enriched ROHs to account for multiple testing.

## Results

### Global and local ancestry analysis in the South African cohort

The SAPDSC exhibited five-way admixture between EUR (56%), AFR (18.8%), NAMA (13%), SAS (6.9%), and MAL (5.2%) (**Supplementary Figure 7**), which is in line with previous studies (Swart *et al*., 2021). The EAS contribution was minimal and the EAS individuals from the 1000 Genomes Project Phase III date were removed from the reference files for downstream analysis, however we retained the MAL individuals as they were shown to contribute notably to LA inference. LA was investigated for each individual in the study to further assess the contribution of ancestral populations at specific genomic regions (**Supplementary Table 5**), allowing for a more precise understanding of the genetic diversity, such as the presence of LD blocks (**Supplementary Figure 8**). It was predicted that the ancestral inference accuracy is as follows (**Supplementary Material**), AFR (∼98.8%), EUR (∼96.5%), MAL (∼82.8%), NAMA (∼86.4%), and SAS (∼86.8%). The ancestral groups with smaller sample sizes and lower-quality genotyping files (SAS, NAMA, and MALAY) have a decreased ancestral inference accuracy. This study serves as a proof of concept for the feasibility of ancestral inference in diverse populations, highlighting the challenges associated with smaller sample sizes and lower-quality data.

### Genome-wide association analysis identifies risk variants associated with Parkinson’s disease

The top hit (rs17098735-T) on chromosome 14 achieved genome-wide significance (beta=2.286; SE=0.401; p-value: 1.23×10^-8^) and is not in LD with any known PD signal. The variant is directly upstream of A-Kinase Anchoring Protein 6 (*AKAP6*) gene, suggesting potential cis-regulatory effects on *AKAP6* transcriptional activity. The encoded protein is highly expressed in various brain regions (**Supplementary Figure 9**), as obtained from the GTEx Analysis Release v10 (dbGaP Accession phs000424.v10.p2) on 01/23/2025 (GTEx Consortium, 2013). Rs17098735-T has a gnomAD frequency of 4.61% in AFR and 0.02% in EUR ancestries. Among the participants carrying the variant (28 cases and 15 controls), 86.7% of the variant carriers were inferred to have AFR ancestry, 11.1% NAMA, and 2.2% MAL ancestry (**Table 1**). No carriers exhibited EUR or SAS inferred ancestry.

**Table 1:** Genome-wide association study top ten hits.

| rsID | Nearest gene(s) | BETA | SE | P-value | AFR<br>LA AF | EUR<br>LA AF | MAL<br>LA AF | NAMA<br>LA AF | SAS<br>LA AF |
| --- | --- | --- | --- | --- | --- | --- | --- | --- | --- |
| rs17098735 | *AKAP6* | 2.286 | 0.401 | $1.23 \times 10^{-8}$ | 86.7% | 0.0% | 2.2% | 11.1% | 0.0% |
| rs562618836 | *RNU6-839P* | -3.901 | 0.728 | $8.45 \times 10^{-8}$ | 96.0% | 0.0% | 0.0% | 4.0% | 0.0% |
| rs190769295 | *ECRG4* | 2.157 | 0.408 | $1.26 \times 10^{-7}$ | 88.9% | 0.0% | 0.0% | 11.1% | 0.0% |
| rs8046041 | *AC093519.2* | 2.436 | 0.475 | $2.90 \times 10^{-7}$ | 84.8% | 0.0% | 0.0% | 15.2% | 0.0% |
| rs75842871 | *KIF18B* | -1.776 | 0.350 | $3.90 \times 10^{-7}$ | 91.0% | 0.0% | 0.0% | 9.0% | 0.0% |
| rs565473607 | *SMNDC1* | -3.694 | 0.741 | $6.12 \times 10^{-7}$ | 91.7% | 0.0% | 4.2% | 4.2% | 0.0% |
| rs140681053 | *RPH3AL* | 2.757 | 0.554 | $6.46 \times 10^{-7}$ | 84.0% | 0.0% | 0.0% | 16.0% | 0.0% |
| rs6490582 | *CRYL1* | 1.267 | 0.256 | $7.69 \times 10^{-7}$ | 16.0% | 59.2% | 3.9% | 14.5% | 6.5% |
| rs58182002 | *CHRNA7* | -3.385 | 0.690 | $9.16 \times 10^{-7}$ | 88.5% | 0.0% | 8.9% | 7.7% | 0.0% |
| rs116582124 | *PREX2* | -2.293 | 0.467 | $9.24 \times 10^{-7}$ | 67.8% | 3.4% | 6.8% | 22.0% | 0.0% |

When looking at suggestive-level variants, one lead locus (rs75842871-G; beta=-1.776; SE=0.350; p-value: 3.90×10^-7^) was found to functionally implicate 17 genes associated with PD (**Supplementary Table 6**) (Simón-Sánchez *et al*., 2009; Do *et al*., 2011; International Parkinson Disease Genomics Consortium *et al*., 2011; Nalls *et al*., 2014; Pickrell *et al*., 2016; Chang *et al*., 2017; Bandres-Ciga *et al*., 2019; Rodrigo and Nyholt, 2021), as identified through Open Target Genetics, which uses a combination of genetic association data, expression quantitative trait loci (eQTL) studies, and functional annotations to link genetic variants to their potential target genes (Ghoussaini *et al*., 2021). We identified one lead locus with a mapped eQTL. This locus, rs75842871-G, was found to be highly expressed in the brain cortex (GTEx Analysis Release v10 (dbGaP Accession phs000424.v10.p2) on 03/05/2025) and significantly associated with Corticotropin Releasing Hormone Receptor 1 (*CRHR1*), with a known PD association (Cheng, Zhu and Zhang, 2020). Additionally, the intergenic variant rs58182002-C (beta=-3.385; SE=0.690; p-value: 9.16×10^-7^) on chromosome 15 influences two genes with an association to PD, Cholinergic Receptor Nicotinic Alpha 7 Subunit (*CHRNA7*) and OTU Deubiquitinase 7A (*OTUD7A*). None of the other top ten lead loci have previous associations with PD or any other neurodegenerative diseases.

The summary statistics and LA information for the top ten GWAS hits are presented in **Table 1**. The Manhattan plot for the GWAS is presented in **Figure 2** and the QQ-plot is presented in **Supplementary Figure 10**. The **λ**GC was 1.113, indicating a slight inflation in the dataset, potentially from using the projected PCA as a covariate in the analysis.

**Figure 2:**
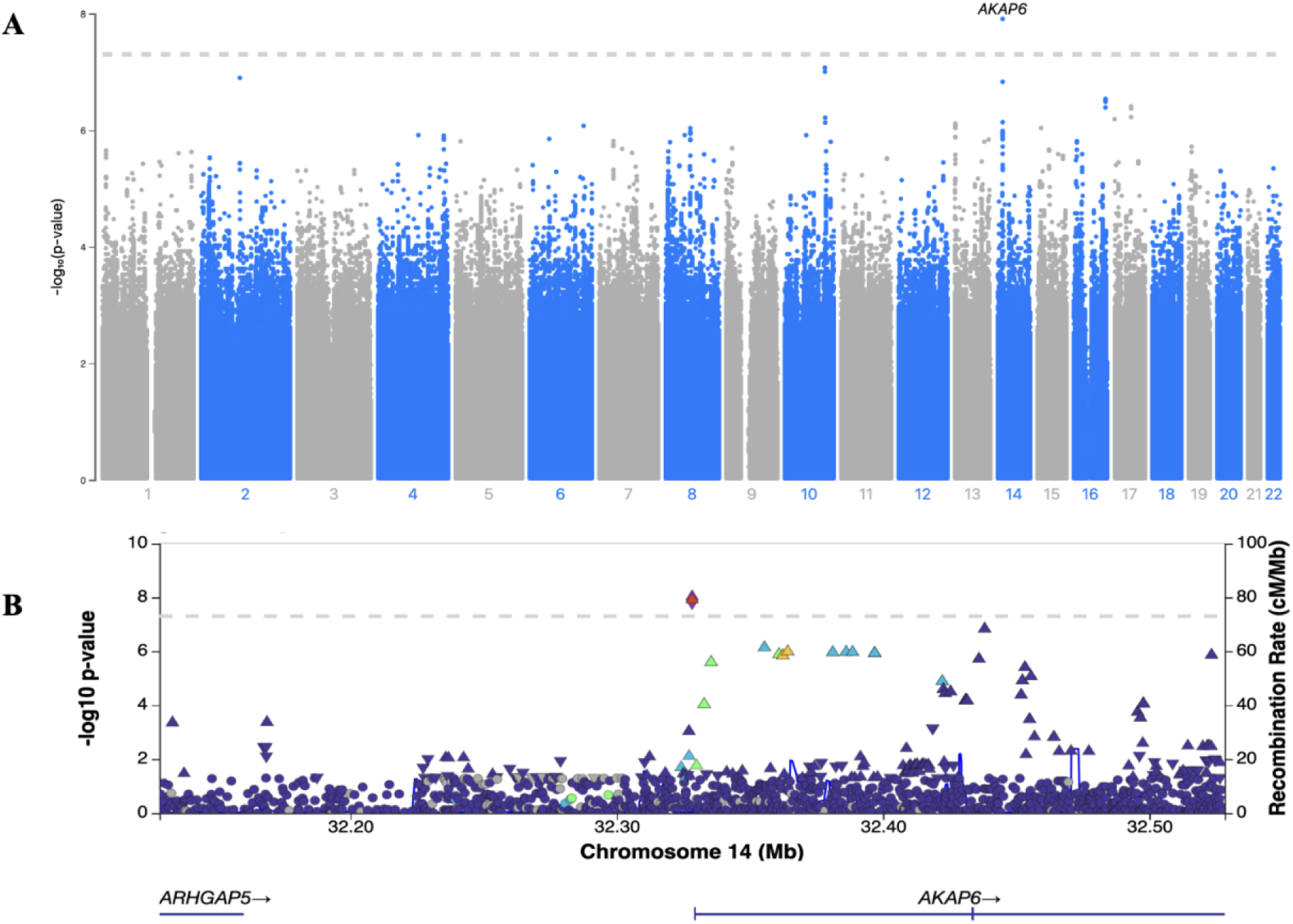
(A) Manhattan plot for SAIGE results by chromosome. The significant peak is located on chromosome 14 within the *AKAP6* gene (B) LocusZoom plot of chromosome 14. The genomic inflation factor (**λ**GC) for p-values ≤0.05 was 1.113.

### Replication and directionality of effect sizes across summary statistics for conventional association study

Although the statistically significant hit (rs17098735-T) was not replicated, three of the suggestively significant lead loci (n=351) were replicated in the LARGE-PD cohort (p-value <0.05), which are located near the Cyclin Dependent Kinase 13 (*CDK13*), Zinc Finger Protein 722 (*ZNF722*), and Calcium And Integrin Binding Family Member 2 (*CIB2*) genes (**Supplementary Table 7**, **Supplementary Figure 11**). Additionally, we examined the p-values for 78 PD GWAS hits identified in Kim *et al*., 2023 and 90 PD GWAS hits identified in Nalls *et al*., 2019, in both the conventional GWAS and LA-GWAS with ATT. In the conventional GWAS (**Supplementary Figure 12**), 11 variants exhibited p-value <0.05 and aligned directionality (Kim *et al*., 2023: n=5, Nalls *et al*., 2019: n=6) and one with opposing directionality (Nalls *et al*., 2019). The LA-GWAS had 18 variants with p-value <0.05 and aligned directionality (Kim *et al*., 2023: n=8, Nalls *et al*., 2019: n=10) and one variant with opposing directionality (Kim *et al*., 2023: n=1). Three of the hits were shared between the conventional GWAS and LA-GWAS, with a total of 28 unique variants with p-value <0.05 between the two methods (**Supplementary Table 8**).

### Local ancestry association analysis reveals additional risk variants

For the LA-GWAS using ATT, there was one lead SNP (rs149066239-T) above genome-wide significance and 35 lead SNPs above the suggestive significance threshold. The top hit (rs149066239-T; beta=-1.826; SE=0.357; p-value: 1.17×10^-8^) is an intergenic variant and eQTL for Amyloid Beta Precursor Protein Binding Family A Member 2 (*APBA2*). Among the loci reaching suggestive significance, we highlight a subset of variants with potential biological relevance. One such variant (rs7601869-T; beta=-1.434; SE=0.295; p-value: 2.51×10^-7^) is an intronic variant nearest to Ubiquitin Conjugating Enzyme E2 E3 (*UBE2E3*), which has been implicated in neurodegenerative disease, including PD (Zheng *et al*., 2016; Mamoor, 2024). Additional variants of interest include rs75614986-G (beta=-1.627; SE=0.350; p-value: 3.14×10^-7^), rs2532570-G (beta=-1.203; SE=0.252; p-value: 5.77×10^-7^), rs139463168-C (beta=-3.120; SE=0.809; p-value: 6.05×10^-7^), and rs10977554-C (beta=0.448; SE=0.092; p-value: 9.29×10^-7^). These variants are associated with genes involved in neuronal pathways, neurodegenerative diseases, and synapse function, making them particularly relevant for further investigation (Kawashima *et al*., 1997; Suzuki *et al*., 2000; Di Gregorio *et al*., 2014; Joo, Hippenmeyer and Luo, 2014; Ishii *et al*., 2022). The LA-GWAS hits were expressed in a range of tissues including the brain, nerve tissue, cultured fibroblasts, tibial nerve, and skeletal muscle ((GTEx Consortium, 2013); dbGaP Accession phs000424.v10.p2). The Manhattan plot for the LA-GWAS using the ATT model is presented in **Figure 3** and the QQ-plot (**λ**GC = 1.311) is presented in **Supplementary Figure 10**. The summary statistics for the top ten lead loci is presented in **Table 2**. One lead locus replicated in the LARGE-PD cohort with aligned directionality (**Supplementary Figure 12**).

**Figure 3:**
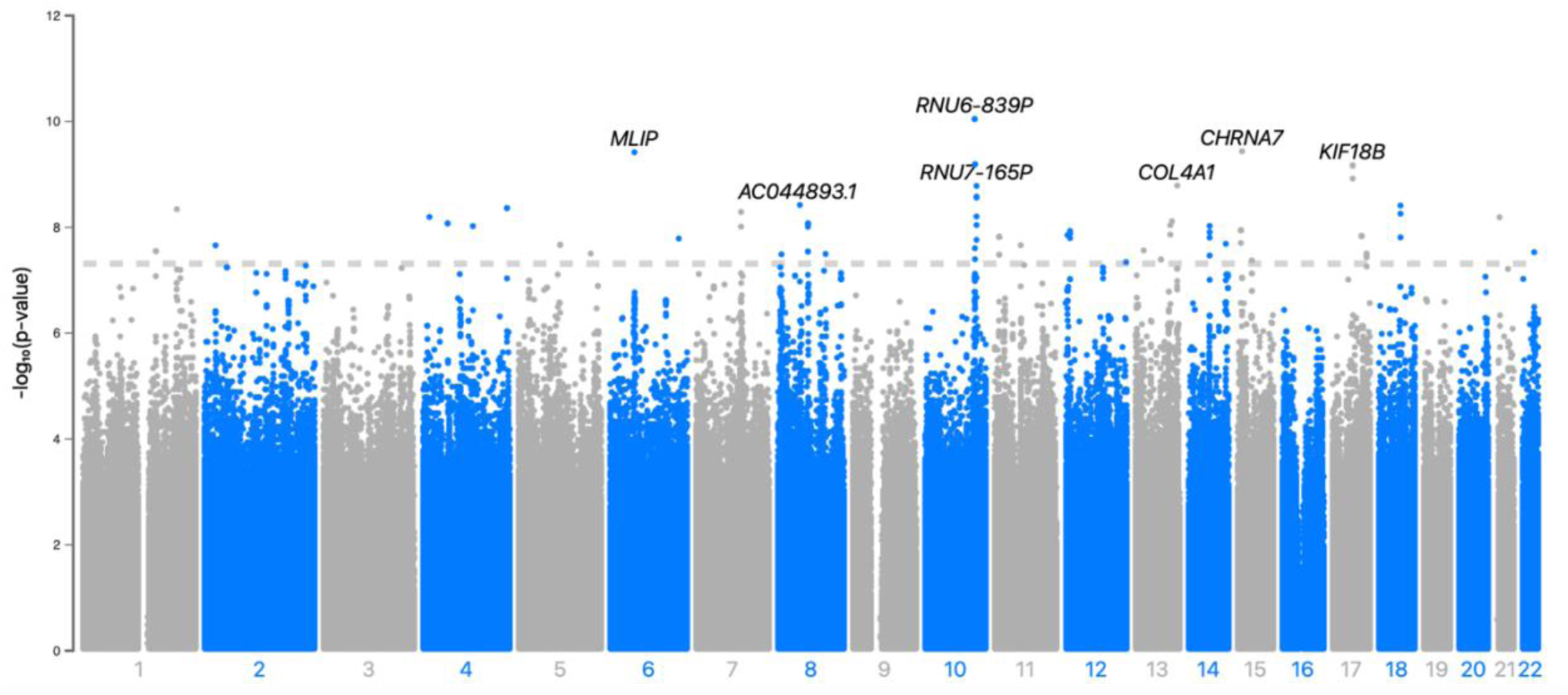
Manhattan plot for LA-GWAS results using ATT model by chromosome. The genomic inflation factor (**λ**GC) for p-values ≤0.05 was 1.311.

**Table 2:**
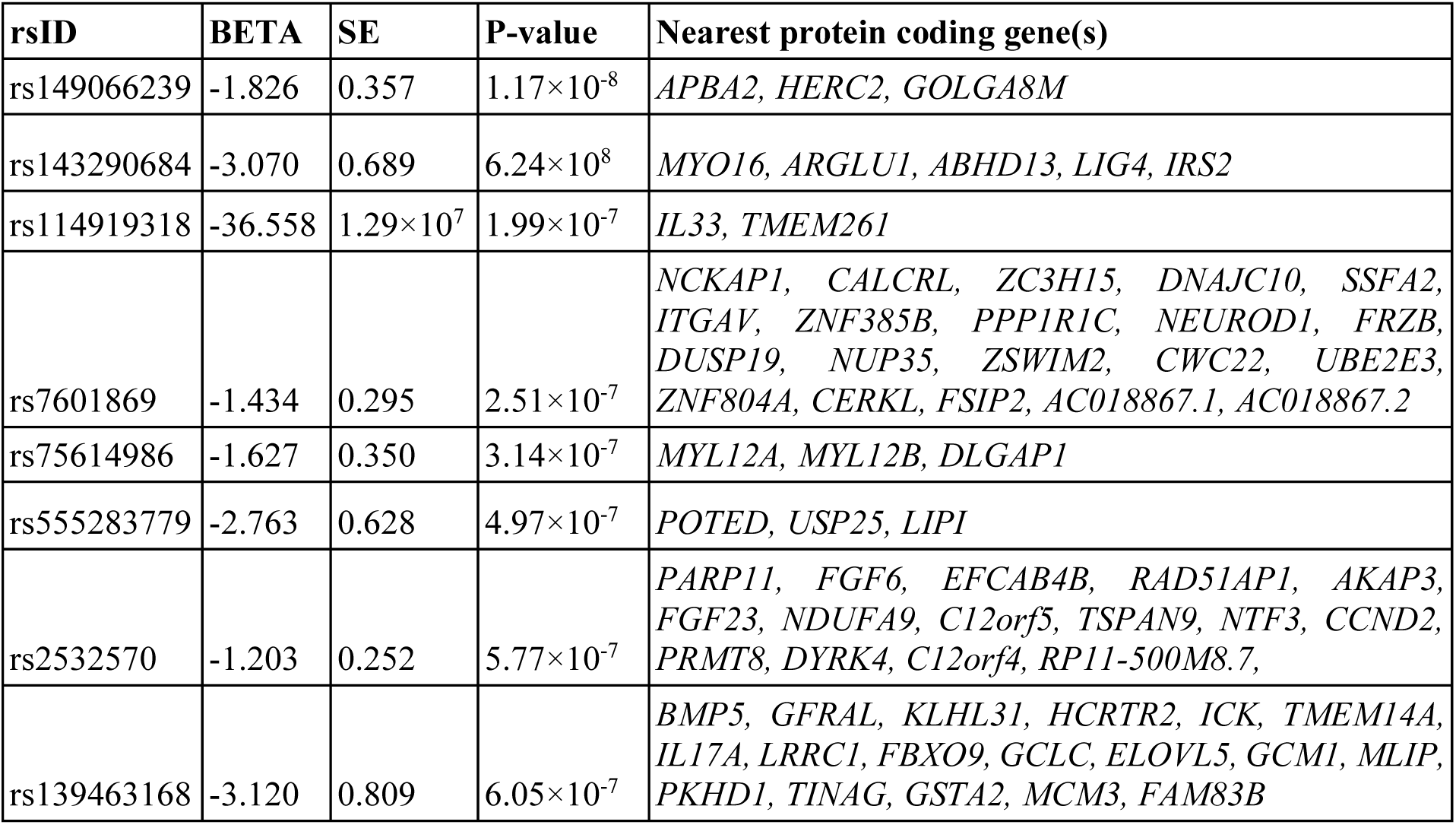
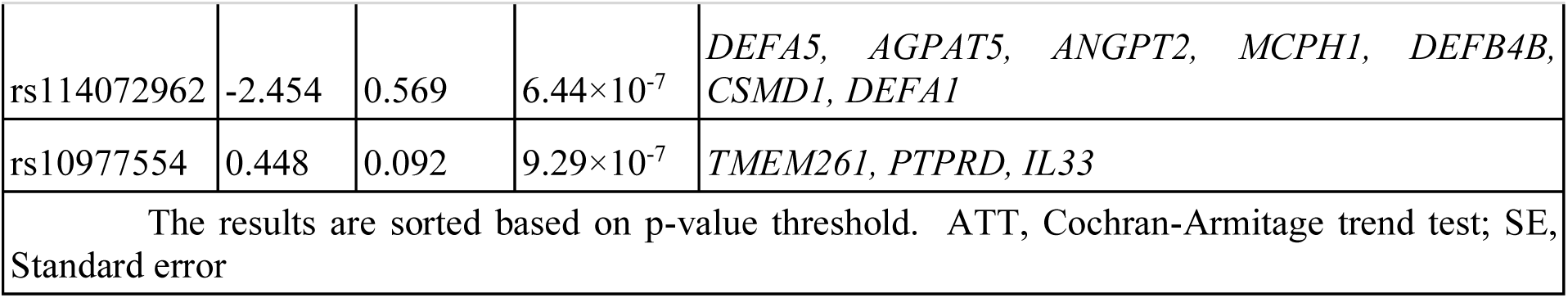
Local ancestry genome-wide association study top ten lead loci for the ATT model.

For the LA-GWAS using Tractor, six lead loci, with suggestive significance, were identified using FUMA (**Supplementary Table 9**), with three lead loci (rs2251850-T (beta=1.480; SE=0.805; p-value: 4.14×10^-7^), rs2449208-T (beta=0.829; SE=1.483; p-value: 7.53×10^-6^), and rs1960758-G (beta=-2.239; SE=1.277; p-value: 9.7×10^-6^)) in the AFR ancestral component and three lead loci (rs4740539-A (beta=1.066; SE=0.277; p-value: 1.31×10^-6^), rs2148353-G (beta=0.985; SE=0.217; p-value:4.84×10^-6^), and rs58553697-G (beta=0.929; SE=0.215; p-value: 7.53×10^-6^)) in all the ancestral components combined (**Figure 4**). No hits reached genome-wide or suggestive significance in the EUR, MAL, NAMA, or SAS ancestries. The top LA-GWAS hit (rs2251850-T), across all ancestries, is an intronic variant which functionally implicates the Wnt Family Member 5A (*WNT5A*) and Leucine Rich Repeats And Transmembrane Domains 1 (*LRTM1*). This lead locus had an allele frequency across the ancestries as follows: AFR (26.6%), EUR (13.6%), MAL (2.1%), NAMA (28.1%), and SAS (11.5%). The lead loci in the AFR component, rs4740539-A, is an intergenic variant for Doublesex And Mab-3 Related Transcription Factors 1, 2, and 3 (*DMRT1*, *DMRT2*, and *DMRT3*). No eQTLs were identified in the LA-GWAS using Tractor.

**Figure 4:**
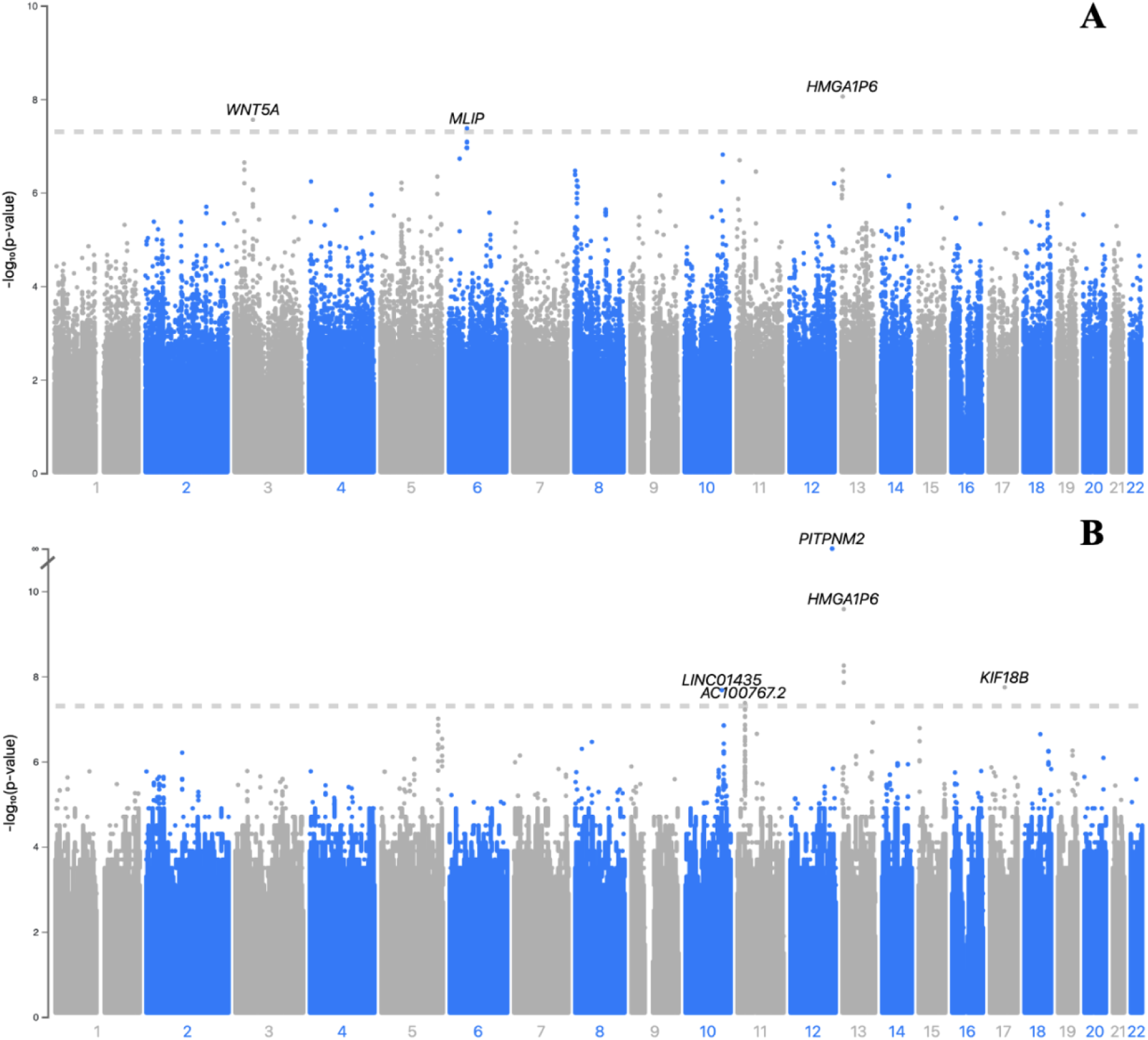
Manhattan plot for LA-GWAS results using Tractor by chromosome for ancestries with genome-wide significance: (A) All ancestries, (B) African. No loci reached genome-wide significance for the European, Nama, Malaysian, or South Asian ancestral components.

### Comparison of runs of homozygosity analysis between the cohorts

A comparison of the ROH analysis results for the SAPDSC and LARGE-PD cohorts is presented in **Table 3**. The total N_ROH_ was 7,087 and 15,501 for the SAPDSC and LARGE-PD cohorts, respectively. The SAPDSC cohort had a lower mean N_ROH_ and S_ROH_ compared to the LARGE-PD cohort (**Supplementary Figure 14**). However, the AV_ROH_ was similar between the two groups. The LARGE-PD cohort showed a higher F_ROH_, indicative of a higher degree of autozygosity suggesting that a greater proportion of this cohort is made up of homozygous segments. Ultimately, there were no statistically significant differences between cases and controls for the four parameters across both cohorts. In terms of investigating ROHs overlapping with known PD, pallido-pyramidal syndrome, and atypical parkinsonism genes and risk loci, LARGE-PD has 443 ROHs overlapping these genes and loci. Among these, 337 ROHs were enriched in cases and three passed Bonferroni correction. Notably, all three ROHs are located on chromosome 7, spanning the same PD region previously identified through GWAS (Nalls *et al*., 2019). Conversely, SAPDSC had 300 overlapping PD genes, 163 enriched in cases, and no ROH that passed Bonferroni correction.

**Table 3:** Runs of homozygosity analysis results.

| Study cohort |  | SAPDSC | LARGE-PD |
| --- | --- | --- | --- |
| NROH | T-test (mean ± SE) | 5.1 ± 0.1 | 10.6 ± 0.2 |
|  | p-value | 0.518 | 0.629 |
| SROH | T-test (mean ± SE) | 13678 ± 503.3 | 32629.9 ± 1192.9 |
|  | p-value | 0.986 | 0.928 |
| AVROH | T-test (mean ± SE) | 2254.8 ± 27.1 | 2676.5 ± 43.7 |
|  | p-value | 0.995 | 0.832 |
| FROH | T-test (mean ± SE) | 0.0046 ± 2×10<sup>-4</sup> | 0.0113 ± 4×10<sup>-4</sup> |
|  | p-value | 986 | 0.928 |
| Number of ROH overlapping PD genes and loci |  | 300 | 443 |
| Number of ROH overlapping PD genes and loci enriched in cases |  | 163 | 337 |
| Number of ROH enriched in cases that pass Bonferroni |  | 0 | 3 |
| Bonferroni correction |  | 3.07×10<sup>-4</sup> | 1.48×10<sup>-4</sup> |
| Legend: AVROH, average length of runs of homozygosity; FROH, runs of homozygosity based estimates for inbreeding coefficient; NROH, number of runs of homozygosity; PD, Parkinson's disease; SAPDSC, South African Parkinson's Disease Study Collection; SE, standard deviation; SROH, total length of runs of homozygosity. p-values indicate the statistical significance as following p < 0.001: ***; p < 0.01: **; p < 0.05: *; p ≥ 0.05: not significant. The Bonferroni threshold was adjusted by dividing 0.05 by the number of enriched ROHs overlapping PD genes to account for multiple comparisons. |  |  |  |

## Discussion

To our knowledge, this study serves as the first PD GWAS including South African participants and the first LA-GWAS for PD in Africa. Ancestry inference revealed the five-way admixture observed in the population, allowing for PD susceptibility to be investigated using population-specific risk variants as well as risk variants for multiple global populations, simultaneously (Swart *et al*., 2021; Step *et al*., 2024).

Our conventional GWAS identified one novel genome-wide significant hit as well as 134 suggestive hits (p-value: 1×10^-5^). The lead locus (rs17098735-T) is located in the *AKAP6* gene and has a CADD v1.7 Phred score of 13.18 indicating it is among the top approximately 5% of most deleterious variants and is likely to have a functional or pathogenic consequence. Proteins encoded by the *AKAP* genes play key roles in regulating numerous brain cellular processes, such as the development, differentiation, and migration of neurons (He *et al*., 2024). The *AKAP6* gene has previously been associated with PD (Phung *et al*., 2020; Pihlstrøm *et al*., 2022; Cappelletti *et al*., 2023). Specifically, differentially methylated loci near *AKAP6* have been linked to the development of Lewy bodies (Pihlstrøm *et al*., 2022) in the brain, a hallmark of PD physiopathology, where reduced *AKAP6* expression has been identified in individuals with PD (Phung *et al*., 2020). Moreover, previous GWAS have identified risk loci in *AKAP6* linked to Alzheimer’s disease (Adewuyi *et al*., 2022), further emphasizing its involvement in the pathogenesis of neurodegenerative diseases. There were numerous hits that reached suggestive significance with potential biological relevance which warrant further study. Many of these are intergenic and intronic variants in genes implicated in various neurological and neurodegenerative disorders, including Alzheimer’s disease (Counts *et al*., 2007) and Parkinson’s disease (Quik, Campos and Grady, 2013).

Of the 135 lead SNPs above the suggestive-significance threshold, three loci replicated in the LARGE-PD cohort, however, this did not include the top GWAS hit. While this number is low, it could be due to population specific risk variants. Additionally, the use of different genotyping arrays between the two cohorts may have contributed to the low replication rate, as differences in array design can affect variant coverage and detection. This is especially true since the MEGA array, used to genotype LARGE-PD Phase 1, did not contain custom content, limiting the ability to identify PD-associated SNPs unless they were common or imputed. Interestingly, the multi-ancestry meta-analysis (Kim *et al*., 2024) consistently showed more variants with aligned directionality. This consistency in effect direction across populations suggests a shared genetic contribution to disease risk, reinforcing the robustness of the association signal across diverse ancestries.

The top hit (rs149066239-T) identified using the LA-GWAS with ATT model is associated with *APBA2* as both an intronic variant and an eQTL. This gene has previously been linked to Alzheimer’s disease (Arnold *et al*., 2013), where the encoded protein acts as a neuronal stabilizer to the amyloid precursor protein, involved in synaptic vesicle exocytosis (Berson *et al*., 2023). One of the lead loci (rs7601869-T) is associated with the protein coding gene, *UBE2E3*, which plays a role in the pathogenesis of neurodegenerative diseases (Zheng *et al*., 2016; Plafker *et al*., 2018). Furthermore, *in silico* prediction shows that this loci has a functional impact on NCK Associated Protein 1 (*NCKAP1*), where the apoptosis of neuronal cells is induced with reduced gene expression (Suzuki *et al*., 2000), and CERK Like Autophagy Regulator (*CERKL*), which plays an essential role in neuronal cell viability (Tuson, Marfany and Gonzàlez-Duarte, 2004).

The LA-GWAS with Tractor identified rs2251850-T, an intergenic variant in *WNT5A* and *LRTM1*, as the top hit across all ancestral components. These proteins are surface markers for midbrain dopaminergic neurons and play a role in their maintenance (Arenas, 2014; Samata *et al*., 2016). Both genes have shown to be potential therapeutic targets for PD treatment (Parish *et al*., 2008; Samata *et al*., 2016). The second (rs2449208-T) and third locus (rs1960758-G) functionally implicate CUB And Sushi Multiple Domains 1 (*CSMD1*), which is highly expressed in the brain (Nagase, Kikuno and Ohara, 2001; Kraus *et al*., 2006). The encoded protein is localised to the synapse (Baum *et al*., 2024) and acts as a regulator for inflammation in the central nervous system (Kraus *et al*., 2006). This gene has previously been implicated in both idiopathic PD (Bai *et al*., 2021) and familial PD (Ruiz-Martínez *et al*., 2017), where pathogenic variants have been absent in neurologically healthy controls.

For the LA-GWAS with Tractor assessing the AFR component, three significant lead loci were identified (rs4740539-A, rs2148353-G, and rs58553697-G). The first and third loci are not in LD and are intergenic variants with *DMRT1*, *DMRT2*, and *DMRT3* respectively. Previous associations with sporadic PD and *DMRT2* have been made in participants of Han Chinese ancestry (Wang *et al*., 2020). Moreover, while *DMRT3* has not been linked directly to PD, it is recognised for its role in gait disturbances (Grillner and El Manira, 2020; Huang *et al*., 2021). The second locus (rs2148353-G) is an intergenic variant for Distal Membrane Arm Assembly Component 1 (*DMAC1*) which is required for a functional accessory subunit required for mitochondrial complex I (Stroud *et al*., 2016). There is significant evidence for the influence of mitochondrial dysfunction, particularly of complex I, in the pathogenesis of PD (Henchcliffe and Beal, 2008; Kamienieva *et al*., 2023; Buck *et al*., 2025). Although no significance was observed in the MAL, NAMA, or SAS ancestry components, this was expected given their minimal representation in the cohort’s inferred ancestry. The lack of significant associations in the EUR group may be due to Tractor distributing genetic risk across multiple ancestry components, reducing the strength of EUR-specific signals in an admixed cohort.

One of the most consistently reported genes in PD GWAS is alpha-synuclein (*SNCA*), as highlighted in previous studies by Nalls *et al*. (2019), Loesch *et al*. (2021), Kim *et al*. (2024), and Foo *et al*. (2020). However, the present study showed no significant peak over the *SNCA* region in the Manhattan plots for any of the GWAS approaches utilized. The *SNCA* SNP closest to significance was rs12648141-G (p-value: 4.4×10^-4^). These observations may reflect a population-specific pattern, suggesting that the role of *SNCA* in PD susceptibility might vary across different ancestral groups. This warrants further investigation, particularly to understand how *SNCA* prevalence and its genetic variations may influence PD risk in AFR and AFR admixed populations.

The ROH analysis found no significant differences between cases and controls in the two study cohorts. However, the mean and SE observed are comparable to previous research investigating the association of PD and ROHs in diverse ancestries, including African and American Admixed cohorts (Step, Hernández, *et al*., 2025). Additionally, the SAPDSC had lower values across all four of the ROH parameters investigated, with shorter and fewer ROHs, suggesting greater admixture in comparison to the LARGE-PD cohort (Ceballos *et al*., 2018). This aligns with previous research showing that while both populations are multi-way admixed, the South African population is five-way admixed (Chimusa *et al*., 2013; Swart *et al*., 2021), while the LARGE-PD cohort is viewed as primarily three-way admixed (Norris *et al*., 2018; Loesch *et al*., 2021).

While we successfully conducted various association analyses, this study has several limitations. The most notable is the small sample size of the SAPDSC. Furthermore, there is no replication dataset for the South African cohort, and we acknowledge two key shortfalls in using LARGE-PD Phase 1 as a replication cohort. First, the datasets were genotyped using different arrays, SAPDSC with the NeuroBooster Array and LARGE-PD Phase 1 using MEGA. Although both arrays provide suitable coverage for imputation, there are differences between them that could impact comparability and coverage. This issue can be rectified using LARGE-PD Phase 2 which uses the NeuroBooster Array, however, data on the complete cohort is not available yet. The second shortfall is the ancestral composition of the replication cohort which differs from the discovery cohort. In LARGE-PD, the proportion of continental AFR ancestry accounts for less than 6% (Loesch *et al*., 2021), making it underpowered for the detection of risk variants specific to participants of AFR ancestry, a major component (18%) of the SAPDSC. Notably, the AFR proportion in LARGE-PD may be even smaller if some individuals have Nama ancestry, a distinct sub-continental population within AFR. Also, both cohorts lack whole-genome sequencing data, limiting further investigation and validation for the ROH analysis. In terms of eQTL mapping, the results generated from FUMA serve as a proof of concept that the GWAS loci function as eQTLs for known PD genes. However, future studies should incorporate RNA sequencing data and a more refined eQTL analysis pipeline to achieve more accurate validation. Finally, while we performed *in silico* validation using public databases, additional *in vitro* functional validation is required to verify the impact of the associated variants on biological pathways.

In conclusion, we provide novel genetic insights into PD pathogenicity using a cohort of highly admixed and genetically diverse South African individuals, highlighting the importance of including genetically diverse and underrepresented populations in PD genetics research. This research aligns with emerging studies demonstrating the value of investigating PD genetics in underrepresented populations (Loesch *et al*., 2021; Rizig *et al*., 2023). These studies have uncovered population-specific variants and mechanisms, collectively underscoring how genetic diversity can reveal previously undetected pathogenic pathways even in modest-sized cohorts.

## Data and code availability

Data used in the preparation of this article were obtained from the Global Parkinson’s Genetics Program (GP2; https://gp2.org). Specifically, we used Tier 2 data from GP2 release 7 [DOI: 10.5281/zenodo.10962119]. Tier 1 data can be accessed by completing a form on the Accelerating Medicines Partnership in Parkinson’s Disease (AMP®-PD) website (https://amp-pd.org/register-for-amp-pd). Tier 2 data access requires approval and a Data Use Agreement signed by your institution. The data analyzed in this study is subject to the following licenses/restrictions: No new genetic data was generated for this study however, summary statistics for the quality and accuracy assessment of the genetic data for the NAMA participants will be made available to researchers who meet the criteria for access after application to the Health Research Ethics Committee of Stellenbosch University. Requests to access the Nama datasets should be directed to Prof. Marlo Moller. Additionally, LARGE-PD summary statistics can be found in the PD GWAS Browser (https://pdgenetics.shinyapps.io/GWASBrowser) and from the Neurodegenerative Disease Knowledge Portal (https://ndkp.hugeamp.org/phenotype.html?phenotype=Parkinsons). The pipelines were developed and are maintained by Dr. Thiago Peixoto Leal and are available at https://github.com/MataLabCCF. Additionally, an overview of the analysis and any additional scripts not available through the Mata Lab GitHub, and the identifiers for all software programs and packages used, are available on GitHub [https://github.com/GP2code/SouthAfrican_PD_GWAS] and were given a persistent identifier via Zenodo [DOI: 10.5281/zenodo.15442537].

## Author contributions

K.S., S.B., and I.M. conceptualized the manuscript. K.S. and T.P.L. wrote the first manuscript draft. T.P.L. compiled the scripts used for the data analysis. T.P.L., E.W., L.M., C.F.H., J.J.F., S.B.-C., and Y.S. assisted with the bioinformatics analysis. All authors reviewed, edited, and approved the final version of the manuscript for submission.

## Acknowledgements

We would like to acknowledge and thank the study participants for their contribution. Data used in the preparation of this article were obtained from Global Parkinson’s Genetics Program (GP2). GP2 is funded by the Aligning Science Across Parkinson’s (ASAP) initiative and implemented by The Michael J. Fox Foundation for Parkinson’s Research (https://gp2.org). For a complete list of GP2 members see https://gp2.org. All figures were created using BioRender (https://www.biorender.com/). We would like to acknowledge Lim Shen-Yang, Tan Ai-Huey, and Azlina Ahmad-Annuar for their efforts in recruiting study participants from Malaysia. We thank Kate Andersh for her work as scientific project manager for this project. We thank Dr. Thiago Peixoto Leal for his contributions to script development used in the analysis. A detailed description of the methods can be found at https://doi.org/10.1101/2025.07.18.25331793. We also acknowledge the Centre for High Performance Computing (CHPC), South Africa, for providing computational resources. We acknowledge the support of the Centre for Tuberculosis Research (CTR) of the South African Medical Research Council. All figures were created using <u>BioRender.com</u>. *For open access, the author has applied a CC BY public copyright license to all Author Accepted Manuscripts arising from this submission*.

