## Supplementary material for "Genome-wide association analyses reveal susceptibility variants linked to Parkinson’s disease in the South African population using inferred global and local ancestry"

Supplementary Figure 2: Box plot to illustrate the distribution of imputation quality across chromosomes generated from the TOPMed Imputation Server. This box plot represents the distribution of empirical R-squared (EmpRsq) values, a measure of imputation accuracy, for each chromosome. EmpRsq is preferred over Rsq for assessing imputation quality because it empirically measures accuracy using observed genotype data, providing a more reliable estimate, especially for rare variants, compared to the model-based Rsq, which can be overly optimistic. The x-axis shows individual chromosomes (CHR), and the y-axis shows EmpRsq values. Higher EmpRsq values suggest more accurate imputation, with an ideal value close to 1. Chromosomes with fewer variants may display a broader distribution. ........................8

Supplementary Figure 3: Principal component analysis for (A) PC1 versus PC2 and (B) PC3 versus PC4. The color dots represent the reference populations, and the black dots represent the South African cohort. AFR, African; EAS, East Asian; MAL, Malaysian; NAMA, Nama-Khoe; SAPDSC, South African Parkinson's Disease Study Collection; SAS, South Asian; PC, Principal component; Pop, population. .9

Supplementary Figure 8: Karyograms showing local ancestry inference for three individuals from the South African Parkinson's Disease Study Collection to illustrate the unique five-way admixture of South African population. Each color represents different ancestral components, highlighting variations in admixture patterns. The karyograms illustrate the distribution of local ancestry blocks along the genome, demonstrating inter-individual differences in ancestral contributions despite shared population background. The ancestries are represented by the following colors: AFR- Purple, EUR- Blue, MAL-Yellow, NAMA- Red, SAS- Orange. chr, chromosome. ........................................................ 12

Supplementary Figure 12: Beta-beta plots of variants identified as significant in previous GWAS studies (Kim *et al*. 2024, Loesch *et al*. 2021, Nalls *et al*. 2019, and Rizig *et al*. 2023), identified as not significant in the SAPDSC conventional GWAS (p-value > 0.05) or nominally significant (p-value < 0.05), generated using ggplot2 (Wickham H *et al*., 2016) package in R (R version 4.4.2 (2024-10-31 ucrt)...16

**Supplementary methods**

### Reference panel information

The reference files followed the same quality control (QC) process as the discovery cohort. After QC, the 1000G, Malaysian (MAL), and Nama (NAMA) ancestry files were merged to create one reference file with all ancestries based on overlapping variants. Reference alignment was confirmed using the Human Reference GRCh38 FASTA file obtained from the Broad Institute, and mismatches were corrected using bcftools v1.9 (Danecek et al. 2021) and the *fixref* plugin. The files were submitted to the TOPMed Imputation Server for phasing using Eagle (Das et al. 2016).

### Power calculation

A priori power analysis was conducted using G*Power v1.3 (Faul et al. 2007) to investigate the reliability of the GWAS results. It is important to note that effect sizes differ between ancestries, and in this analysis, we used the African cohort odds ratio of 1.58 (Rizig et al. 2023).

The following input was used to calculate the sample size to achieve 80% power.
Test family: Z test
Statistical test: Logistic regression
Type of power analysis: A priori: Compute required sample size- give α, power, and effect size
Input parameters:

- Tail(s): one
- Odds ratio: 1.58 (Rizig et al. 2023)
- Pr(Y=1|X=1) H0: 0.3
- A err prob: 0.05
- Power (1-β err prob): 0.8
- $R^2$ other X: 0
- X distribution: Normal
- X parm μ: 0
- X parm σ: 1

The a priori power analysis determined that a sample size of 151 participants would provide 80% power to detect the expected effect size, with a critical z-value of 1.64.

### Ancestry inference accuracy calculation

To calculate the accuracy of the prediction based on the confusion matrix output, use the following formula:

Accuracy= $\frac{\Sigma\ diagonal\ elements}{\Sigma\ all\ elements}$

### Polymerase chain reaction and Sanger sequencing

To confirm the presence of the top GWAS hit, Sanger sequencing was performed for all variant carriers (n=43). Polymerase chain reaction (PCR) primers were designed using Primer Quest software (https://www.idtdna.com/SciTools; Assessed: 04 June 2025). The forward primer sequence is 5' TAG GCT GCT CTG TCC ATA TCT 3' and the reverse primer sequence is 5' ACT GCC TAC CAC ACA GAT TAT T 3'. The PCR product was 210 base pairs in length. The following reagents and quantities were used: thermoscientific DreamTaq™ Hot Start PCR Master Mix (25µl), $dH_2O$ (22µl), forward primer (1µl), reverse primer (1µl), with 1µl DNA or non-template control. The PCR was performed using the GeneAmp PCR System 2700 (Applied Biosystems Forster City, USA). The PCR conditions were as follows: initial denaturation at 95℃, 30 cycles at 30 seconds each of denaturation at 95℃, annealing at 58℃, extension at 72℃, and a final extension at 72℃ for 10 minutes. The PCR products were viewed using a 2% agarose gel electrophoresis. Sanger sequencing was performed using the BigDye Terminator V3.1 sequencing kit (Applied Biosystems, USA) according to the manufacturer's instructions at the Central Analytical Facilities (CAF) at Stellenbosch University, South Africa.

**Supplementary figures**

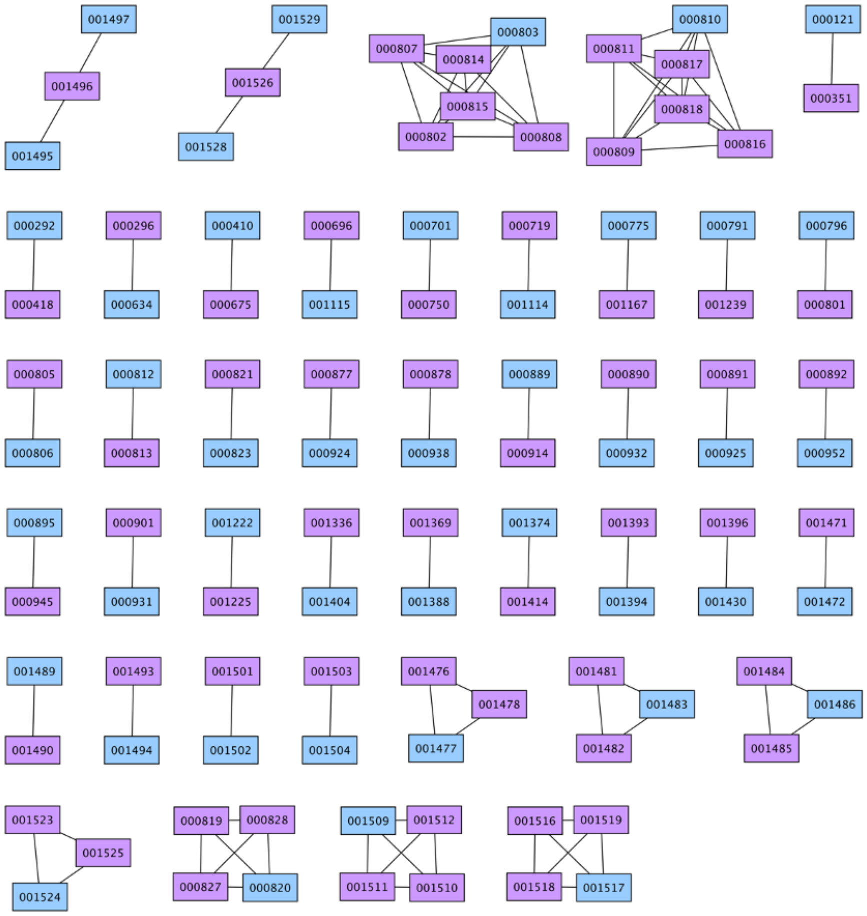

**Supplementary Figure 1:** Relationship inference using node centrality method. The relationship and family structures are indicated with the lines. The individuals in purple were removed for the analysis, while the individuals in blue were retained.

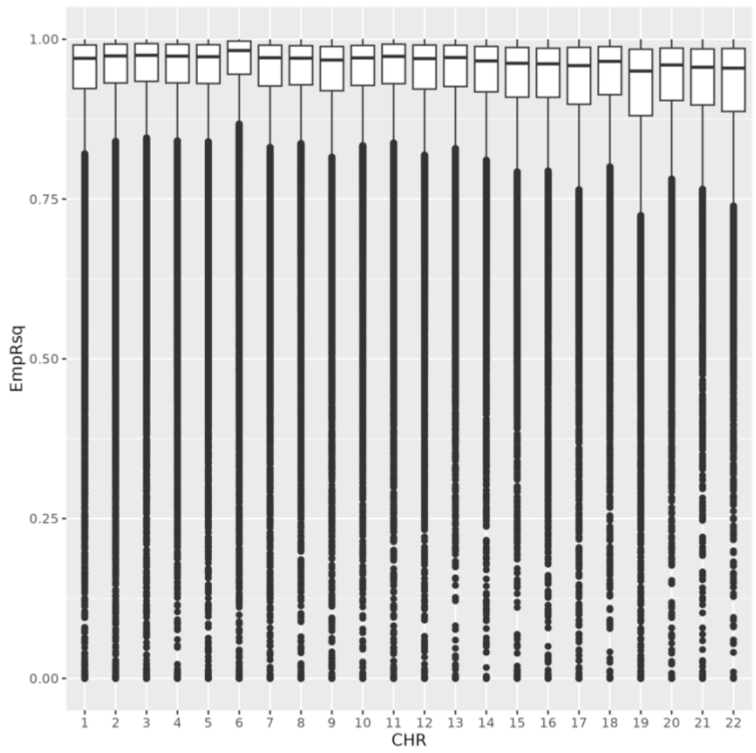

**Supplementary Figure 2:** Box plot to illustrate the distribution of imputation quality across chromosomes generated from the TOPMed Imputation Server. This box plot represents the distribution of empirical R-squared (EmpRsq) values, a measure of imputation accuracy, for each chromosome. EmpRsq is preferred over Rsq for assessing imputation quality because it empirically measures accuracy using observed genotype data, providing a more reliable estimate, especially for rare variants, compared to the model-based Rsq, which can be overly optimistic. The x-axis shows individual chromosomes (CHR), and the y-axis shows EmpRsq values. Higher EmpRsq values suggest more accurate imputation, with an ideal value close to 1. Chromosomes with fewer variants may display a broader distribution.

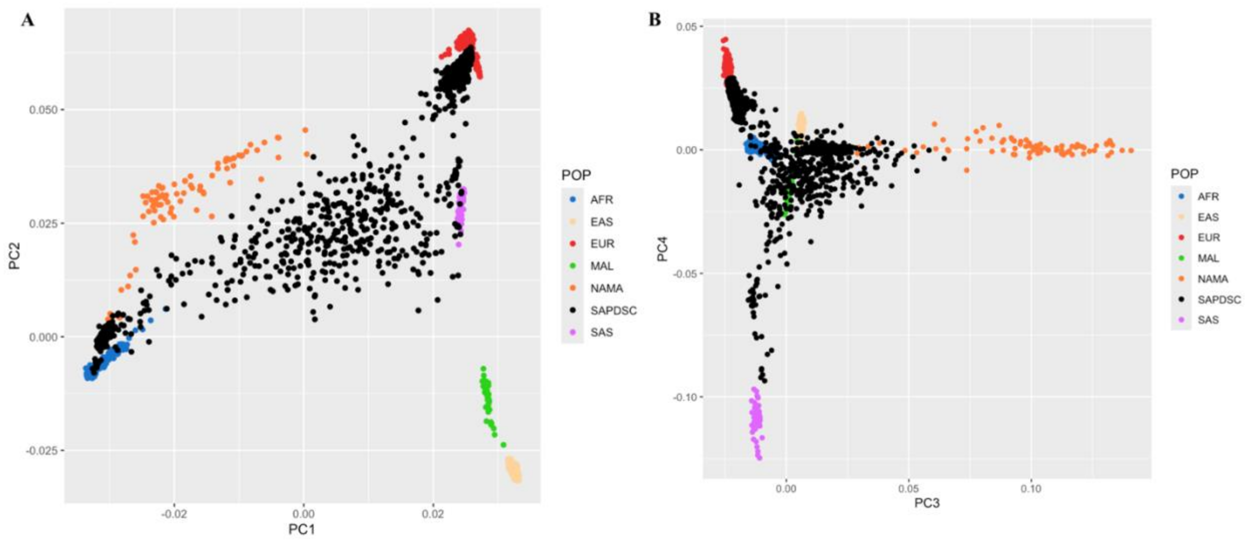

**Supplementary Figure 3:** Principal component analysis for (A) PC1 versus PC2 and (B) PC3 versus PC4. The color dots represent the reference populations, and the black dots represent the South African cohort. AFR, African; EAS, East Asian; MAL, Malaysian; NAMA, Nama-Khoe; SAPDSC, South African Parkinson's Disease Study Collection; SAS, South Asian; PC, Principal component; Pop, population.

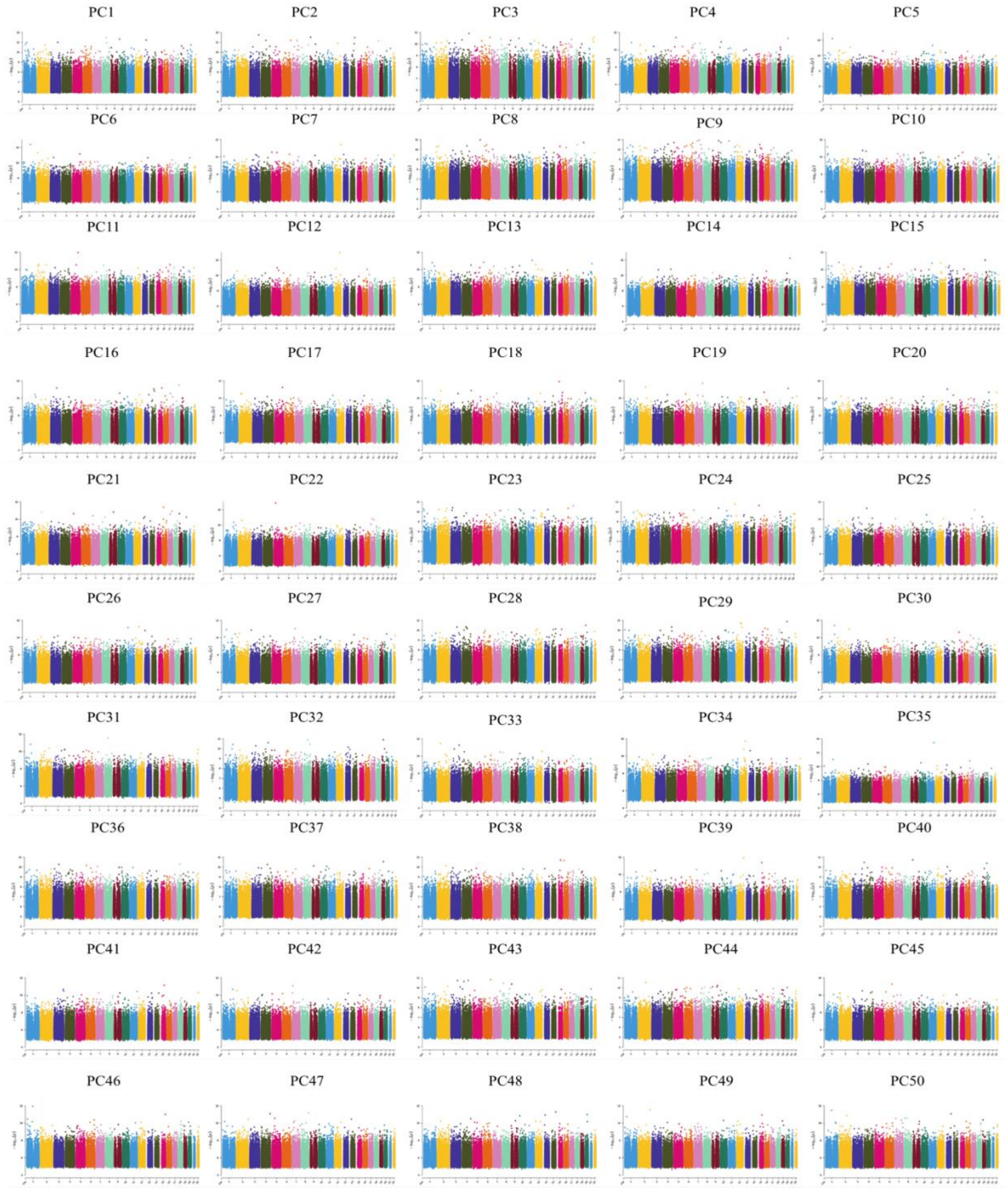

**Supplementary Figure 4:** Manhattan plots to assess the genome-wide influence of the Principal Components 1-50. PC, Principal component.

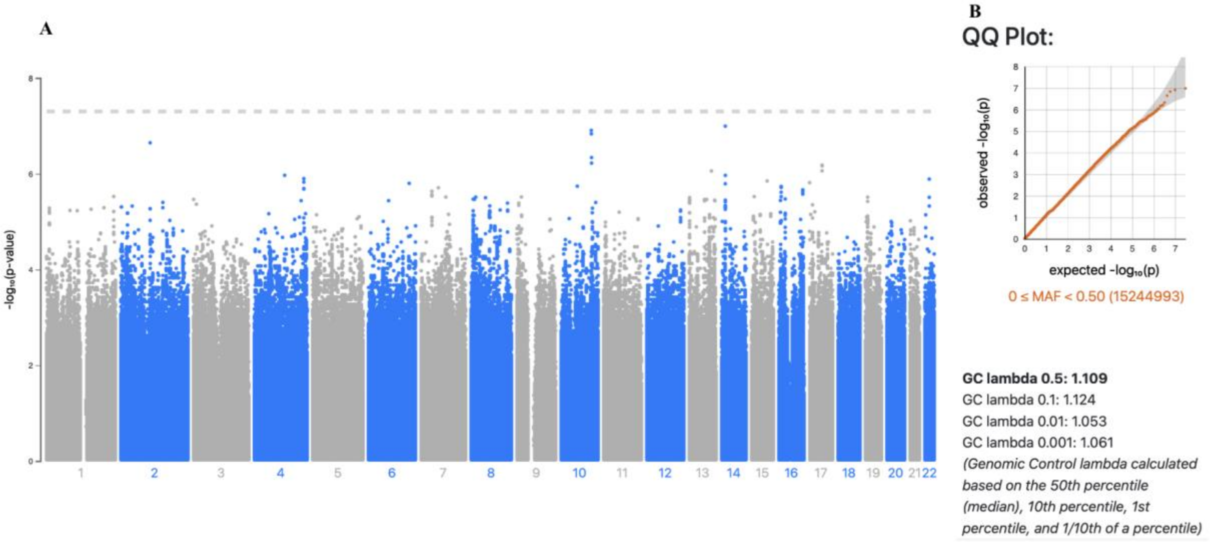

**Supplementary Figure 5:** (A) Manhattan plot for the association results using Principal components 1-10. (B) QQ-plot of the GWAS p-values and GC lambda values.

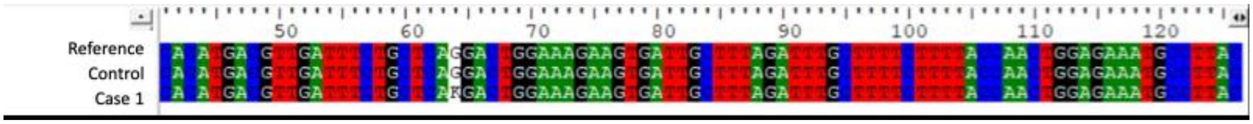

**Supplementary Figure 6:** Sequencing chromatograms to confirm the presence of the top hit identified through the conventional genome-wide association study.

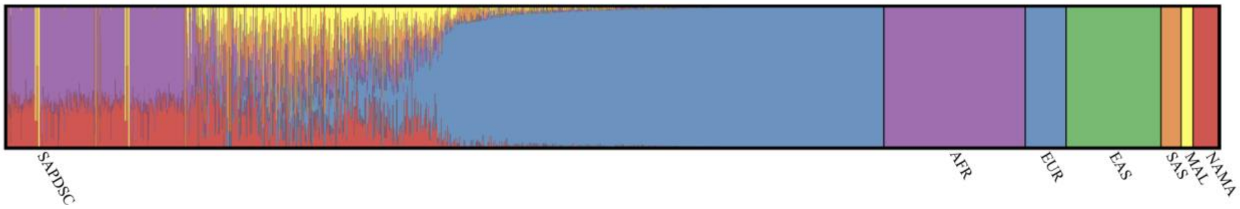

**Supplementary Figure 7:** Forest plot generated after running ADMIXTURE (k=6) using pong software. Each vertical line represents one individual and each color represents an ancestry. AFR, African; EAS, East Asian; EUR, European; MAL, Malaysian; NAMA, Nama-Khoe; SAPDSC, South African Parkinson's Disease Study Collection; SAS, South Asian

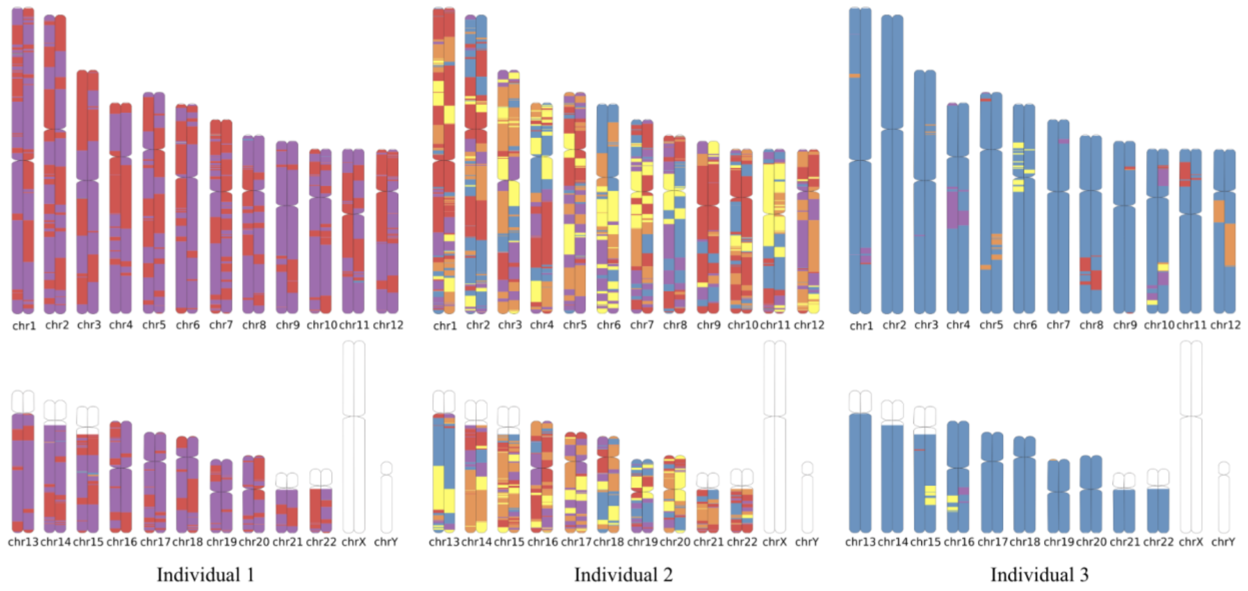

**Supplementary Figure 8:** Karyograms showing local ancestry inference for three individuals from the South African Parkinson's Disease Study Collection to illustrate the unique five-way admixture of South African population. Each color represents different ancestral components, highlighting variations in admixture patterns. The karyograms illustrate the distribution of local ancestry blocks along the genome, demonstrating inter-individual differences in ancestral contributions despite shared population background. The ancestries are represented by the following colors: AFR- Purple, EUR- Blue, MAL- Yellow, NAMA- Red, SAS- Orange. chr, chromosome.

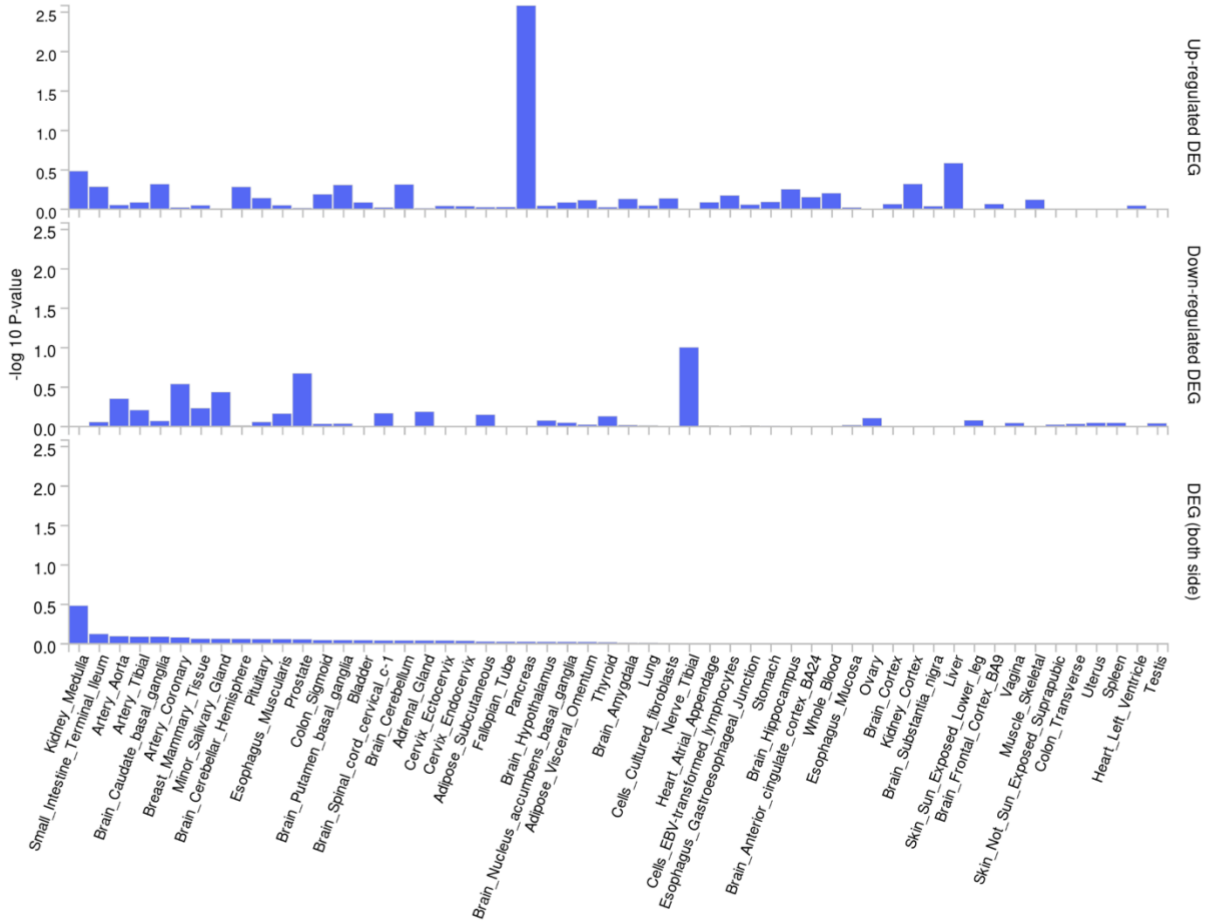

**Supplementary Figure 9:** GTEx-based plot showing the association between genetic variants and gene expression across 54 tissue types, derived from the SAPDSC GWAS summary statistics. GWAS, genome-wide association study; SAPDSC, South African Parkinson's Disease Study Collection

A

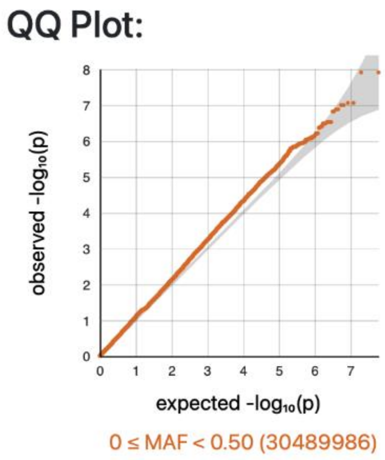

**GC lambda 0.5: 1.113**
GC lambda 0.1: 1.127
GC lambda 0.01: 1.074
GC lambda 0.001: 1.093
*(Genomic Control lambda calculated based on the 50th percentile (median), 10th percentile, 1st percentile, and 1/10th of a percentile)*

B

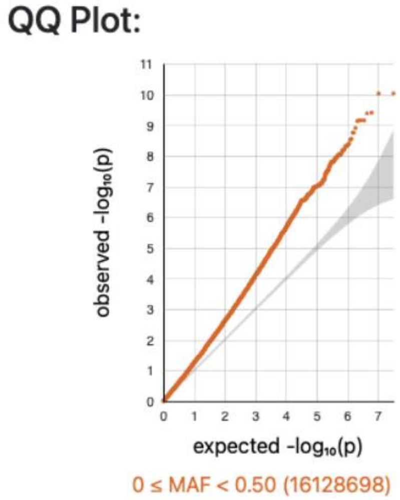

**GC lambda 0.5: 1.311**
GC lambda 0.1: 1.332
GC lambda 0.01: 1.378
GC lambda 0.001: 1.433
*(Genomic Control lambda calculated based on the 50th percentile (median), 10th percentile, 1st percentile, and 1/10th of a percentile)*

**Supplementary Figure 10:** QQ-plot recommended principal components from (A) step-wise regression as well as the GC lambda values. (B) QQ-plot for the LA-GWAS using ATT as well as the GC lambda values. GC, Genomic control; QQ, Quantile-Quantile

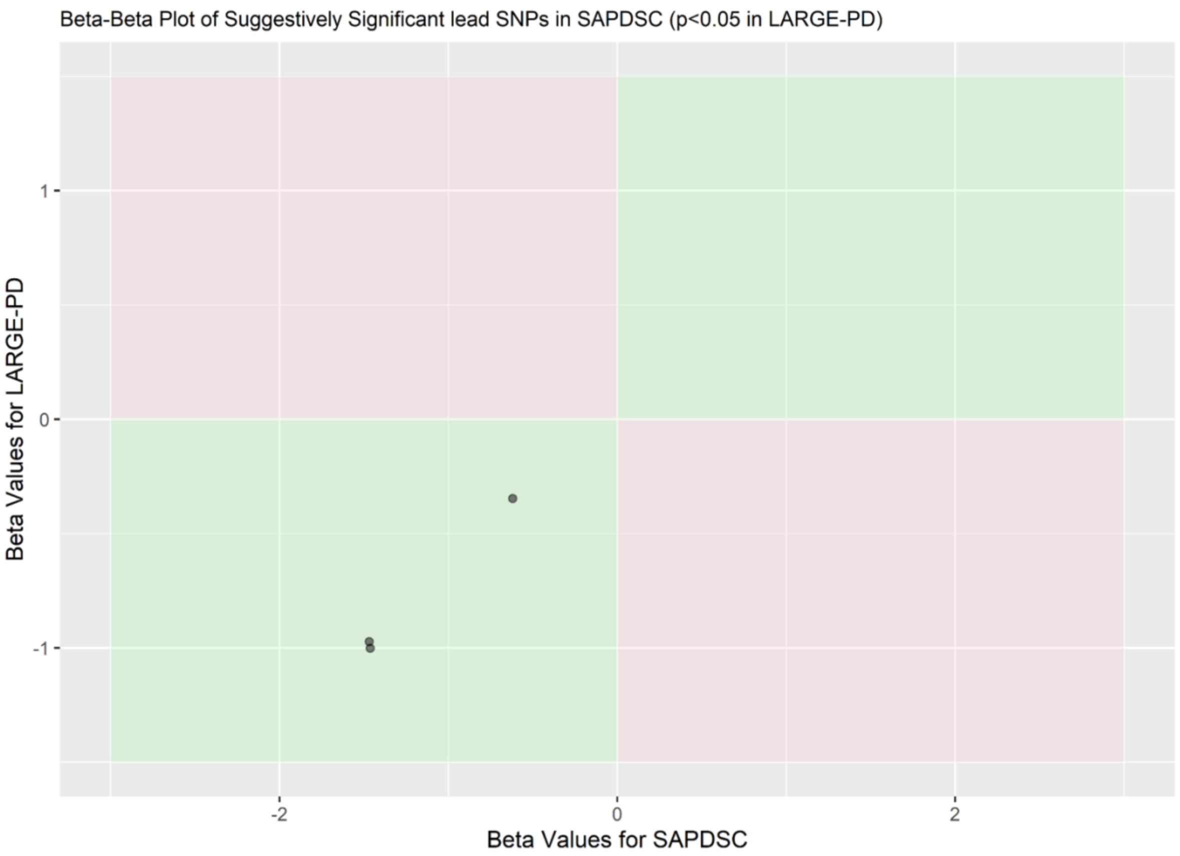

**Supplementary Figure 11:** Beta-beta plots for replication of variants identified as suggestively significant in the SAPDSC conventional GWAS compared to those with p-value < 0.05 in the LARGE-PD replication cohort, generated using ggplot2 (Wickham H *et al*., 2016) package in R (R version 4.4.2 (2024-10-31 ucrt).

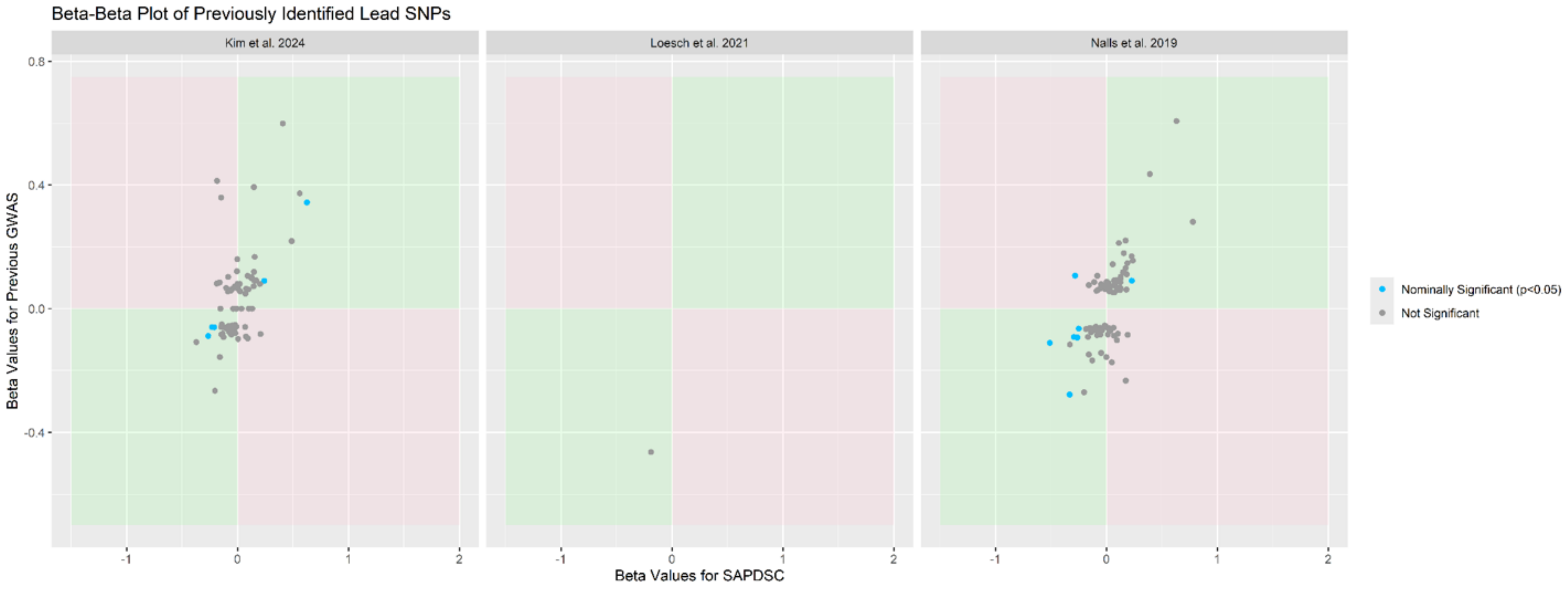

**Supplementary Figure 12:** Beta-beta plots of variants identified as significant in previous GWAS studies (Kim *et al*. 2024, Loesch *et al*. 2021, Nalls *et al*. 2019, and Rizig *et al*. 2023), identified as not significant in the SAPDSC conventional GWAS (p-value > 0.05) or nominally significant (p-value < 0.05), generated using ggplot2 (Wickham H *et al*., 2016) package in R (R version 4.4.2 (2024-10-31 ucrt).

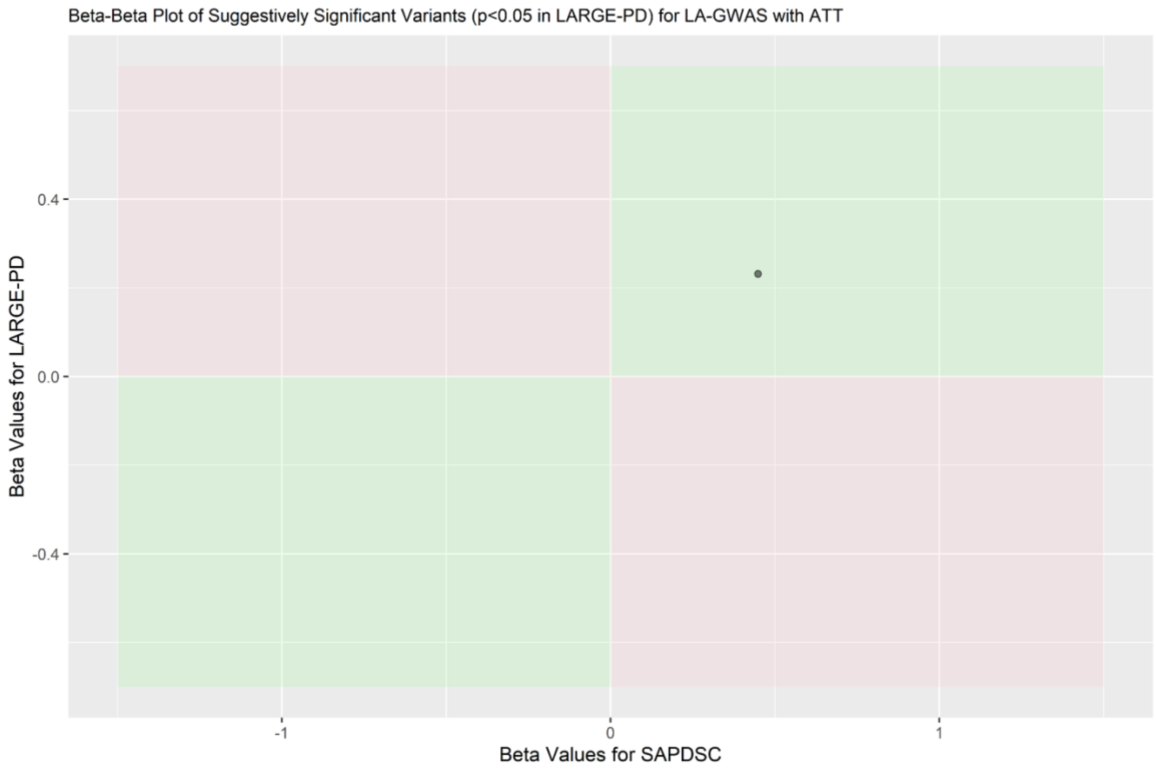

**Supplementary Figure 13:** Beta-beta plots of the lead loci in the SAPDSC local ancestry association separated with ATT, as well as had p-values < 0.05 in Loesch *et al*. 2021, generated using ggplot2 (Wickham H *et al*., 2016) package in R (R version 4.4.2 (2024-10-31 ucrt). p, p-value; SAPDSC, South African Parkinson's Disease Study Collection.

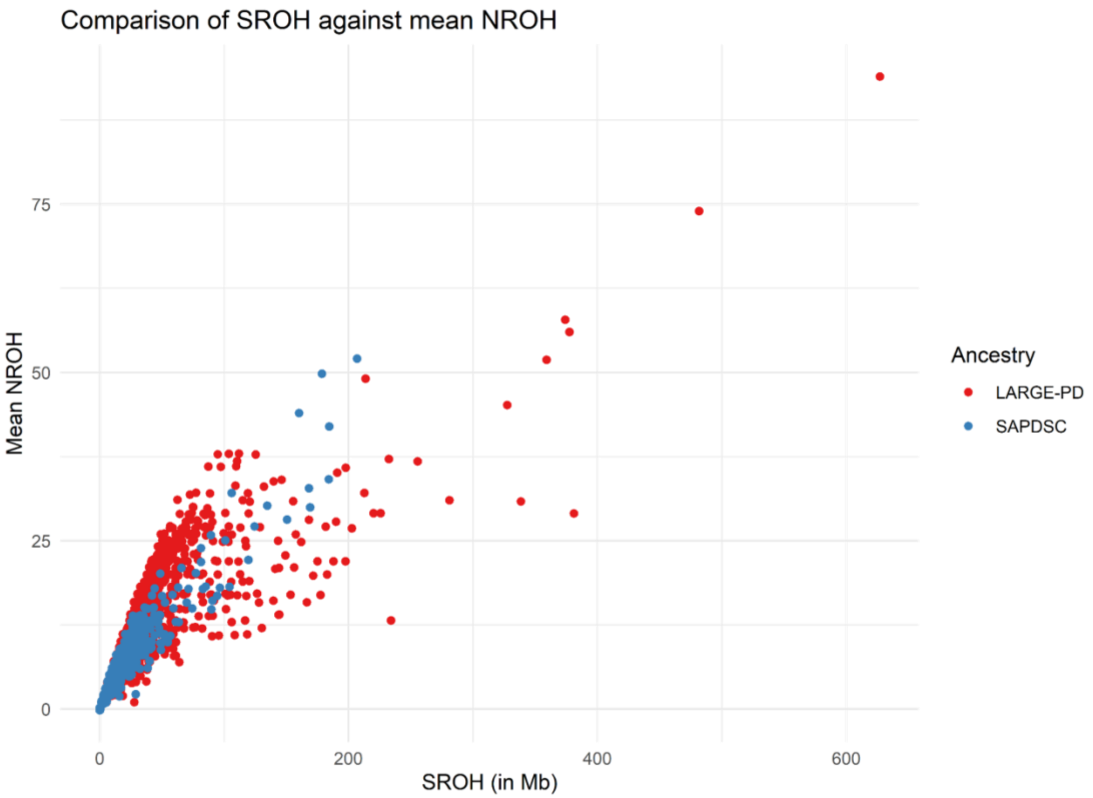

**Supplementary Figure 14:** Comparison for the mean number of runs of homozygosity and the sum of the number of runs of homozygosity for the LARGE-PD and SAPDSC. LPD, Latin American Research Consortium on the Genetics of Parkinson’s Disease; Mb, megabases; NROH, number of runs of homozygosity; SAPDSC, South African Parkinson’s Disease Study Collection; SROH, sum of the runs of homozygosity.

**Supplementary tables**

**Supplementary Table 1:** South African Parkinson's Disease Study Collection demographic information

| | Total participants | Number of males | Number of females | Mean (SD) age | Mean (SD) AAO |
|---|---|---|---|---|---|
| **PD Cases** | 691 | 398 (57.60%) | 293 (42.40%) | 65.77 (12.06) | 57.44 (12.56) |
| **Controls** | 826 | 385 (46.61%) | 441 (53.39%) | 44.72 (21.49) | N/A |
| **Total** | 1517 | 783 (51.62%) | 734 (48.38%) | 54.31 (20.67) | N/A |
| Legend: AAO, Age at onset; N/A, Not applicable; SD, standard deviation | | | | | |

**Supplementary Table 2:** Replication cohort sample size, sex, and disease status by recruitment institution

| Country | Institution | Total participants | PD cases | Controls | Number of males | Number of females | Mean (SD) age |
|---|---|---|---|---|---|---|---|
| **Brazil** | Hospital de Clínicas de Porto Alegre | 13 | 3 | 10 | 2 | 11 | 64.46 (9.12) |
| | Hospital das Clínicas da Faculdade de Medicina de Ribeirão Preto | 195 | 126 | 69 | 96 | 99 | 57.15 (15.12) |
| | Hospital das Clínicas da Faculdade de Medicina da Universidade de São Paulo | 19 | 19 | 0 | 14 | 5 | 59.37 (9.04) |
| **Chile** | Centro de Trastornos del Movimiento (CETRAM) | 13 | 13 | 0 | 10 | 3 | 63.67 (9.59) |
| **Colombia** | Universidad Nacional de Colombia | 23 | 23 | 0 | 9 | 14 | 49.09 (10.18) |
| | Universidad de Antioquia | 328 | 134 | 194 | 143 | 185 | 60.34 (12.44) |
| **Peru** | Instituto Nacional de Ciencias Neurológicas | 670 | 437 | 233 | 307 | 363 | 60.15 (13.57) |
| | Universidad Nacional del Altiplano | 45 | 0 | 45 | 11 | 34 | 43.00 (19.80) |

| | | | | | | | |
|---|---|---|---|---|---|---|---|
| **Uruguay** | Universidad de la República | 191 | 52 | 139 | 67 | 124 | 61.13 (12.62) |
| **TOTAL** | | 1497 | 807 | 690 | 659 | 838 | 59.30 (13.90) |
| Legend: PD, Parkinson's disease; SD, Standard deviation | | | | | | | |

**Supplementary Table 3:** Demographics for reference files

| **Source** | **Population** | **Number of participants** |
|---|---|---|
| 1000 Genomes | African | 471 |
| 1000 Genomes | East Asian | 316 |
| 1000 Genomes | European | 136 |
| 1000 Genomes | South Asian | 67 |
| Global Parkinson's Genetics Program | Malaysian | 40 |
| Tuberculosis Host Genetics Research Group Stellenbosch University | Nama | 84 |

**Supplementary Table 4:** List of the known Parkinson's disease, pallido-pyramidal syndrome, and atypical parkinsonism gene regions and risk loci

| **Marker name** | **Near gene** | **Source** |
|---|---|---|
| chr5:75803383-76803383 | *SV2C* | (Foo et al., 2020) |
| chr7:70785507-71785507 | *GALNT17* | (Foo et al., 2020) |
| chr22:49869588-50869588 | *PPP6R2* | (Kim et al., 2024) |
| chr13:109310821-110310821 | *RN7SKP10* | (Kim et al., 2024) |
| chr1:92586630-93586630 | *MTF2* | (Kim et al., 2024) |
| chr17:15830946-16830946 | *PIGL* | (Kim et al., 2024) |
| chr4:2347441-3347441 | *ADD1* | (Kim et al., 2024) |
| chr21:14940233-15940233 | *AJ009632.2* | (Kim et al., 2024) |
| chr15:49995212-50995212 | *USP8* | (Kim et al., 2024) |
| chr17:81587210-82587210 | *FASN* | (Kim et al., 2024) |
| chr20:31333121-32333121 | *MYLK2* | (Kim et al., 2024) |
| chr8:109132545-110132545 | *SYBU* | (Kim et al., 2024) |
| chr3:178643629-179643629 | *AC076966.1* | (Kim et al., 2024) |
| chr22:40538154-41538154 | *RBX1* | (Kim et al., 2024) |
| chr16:52435514-53435514 | *PHBP2* | (Nalls et al., 2019) |
| chr3:182542285-183542285 | *MCCC1* | (Nalls et al., 2019) |
| chr10:101755522-102755522 | *GBF1* | (Nalls et al. 2019) |

| | | |
|---|---|---|
| chr9:17227067-18227067 | *SH3GL2* | (Nalls et al. 2019) |
| chr1:232028865-233028865 | *SIPA1L2* | (Nalls et al. 2019) |
| chr12:122342051-123342051 | *HIP1R* | (Nalls et al. 2019) |
| chr16:30466478-31466478 | *SETD1A* | (Nalls et al. 2019) |
| chr14:54382151-55382151 | *GCH1* | (Nalls et al. 2019) |
| chr6:32110995-33110995 | *AHR* | (Nalls et al. 2019) |
| chr1:154425709-155425709 | *PMVK* | (Nalls et al. 2019) |
| chr1:205268611-206268611 | *RAB29* | (Nalls et al. 2019) |
| chr1:171250629-172250629 | *VAMP4* | (Nalls et al. 2019) |
| chr12:131987182-132987182 | *FBRSL1* | (Nalls et al. 2019) |
| chr17:46289439-47289439 | *WNT3* | (Nalls et al. 2019) |
| chr2:101280501-102280501 | *MAP4K4* | (Nalls et al. 2019) |
| chr3:150891177-151891177 | *MED12L* | (Nalls et al. 2019) |
| chr17:45220942-46220942 | *MAPT* | (Nalls et al. 2019) |
| chr10:119276815-120276815 | *INPP5F* | (Nalls et al. 2019) |
| chr5:134363415-135363415 | *AC006077.1* | (Nalls et al. 2019) |
| chr14:37020065-38020065 | *MIPOL1* | (Nalls et al. 2019) |
| chr11:83276234-84276234 | *DLG2* | (Nalls et al. 2019) |
| chr18:42593415-43593415 | *RIT2* | (Nalls et al. 2019) |
| chr3:48211556-49211556 | *IP6K2* | (Nalls et al. 2019) |
| chr6:71278059-72278059 | *RIMS1* | (Nalls et al. 2019) |
| chr17:6952302-7952302 | *MAPT* | (Nalls et al. 2019) |
| chr8:11354934-12354934 | *CTSB* | (Nalls et al. 2019) |
| chr17:42088995-43088995 | *RETREG3* | (Nalls et al. 2019) |
| chr4:112947909-113947909 | *CAMK2D* | (Nalls et al. 2019) |
| chr9:17079692-18079692 | *SH3GL2* | (Nalls et al. 2019) |
| chr3:160859842-161859842 | *SPTSSB* | (Nalls et al. 2019) |
| chr2:167753884-168753884 | *STK39* | (Nalls et al. 2019) |
| chr5:60342132-61342132 | *ELOVL7* | (Nalls et al. 2019) |
| chr18:33224354-34224354 | *ASXL3* | (Nalls et al. 2019) |
| chr7:22760430-23760430 | *GPNMB* | (Nalls et al. 2019) |
| chr2:94835195-95835195 | *KCNIP3* | (Nalls et al. 2019) |
| chr8:129389663-130389663 | *FAM49B* | (Nalls et al. 2019) |
| chr21:36980059-37980059 | *DYRK1A* | (Nalls et al. 2019) |
| chr15:61205186-62205186 | *lincRNA* | (Nalls et al. 2019) |
| chr17:43716969-44716969 | *UBTF* | (Nalls et al. 2019) |
| chr8:22168467-23168467 | *BIN3* | (Nalls et al. 2019) |
| chr5:102530090-103530090 | *PAM* | (Nalls et al. 2019) |
| chr16:28433075-29433075 | *CD19* | (Nalls et al. 2019) |

| | | |
|---|---|---|
| chr16:52102330-53102330 | *CASC16* | (Nalls et al. 2019) |
| chr4:17467188-18467188 | *LCORL* | (Nalls et al. 2019) |
| chr4:458159-1458159 | *TMEM175* | (Nalls et al. 2019) |
| chr12:39840400-40840400 | *LRRK2* | (Nalls et al. 2019) |
| chr4:89204960-90204960 | *AC097478.1* | (Nalls et al. 2019) |
| chr1:154662560-155662560 | *KRTCAP2* | (Nalls et al. 2019) |
| chr14:74406331-75406331 | *RPS6KL1* | (Nalls et al. 2019) |
| chr11:133417106-134417106 | *IGSF9B* | (Nalls et al. 2019) |
| chr4:75726816-76726816 | *FAM47E* | (Nalls et al. 2019) |
| chr6:27271022-28271022 | *RSL24D1P1* | (Nalls et al. 2019) |
| chr1:226228377-227228377 | *ITPKB* | (Nalls et al. 2019) |
| chr4:15235725-16235725 | *BST1* | (Nalls et al. 2019) |
| chr13:96712767-97712767 | *MBNL2* | (Nalls et al. 2019) |
| chr4:89215479-90215479 | *AC097478.1* | (Nalls et al. 2019) |
| chr19:1841049-2841049 | *SPPL2B* | (Nalls et al. 2019) |
| chr3:121978045-122978045 | *KPNA1* | (Nalls et al. 2019) |
| chr2:134207046-135207046 | *TMEM163* | (Nalls et al. 2019) |
| chr17:61340005-62340005 | *BRIP1* | (Nalls et al. 2019) |
| chr8:16340084-17340084 | *AC011586.2* | (Nalls et al. 2019) |
| chr17:45166837-46166837 | *MAPT* | (Nalls et al. 2019) |
| chr4:169162006-170162006 | *CLCN3* | (Nalls et al. 2019) |
| chr9:33546393-34546393 | *UBAP2* | (Nalls et al. 2019) |
| chr16:18766171-19766171 | *SYT17* | (Nalls et al. 2019) |
| chr16:50202745-51202745 | *NOD2* | (Nalls et al. 2019) |
| chr1:160999264-161999264 | *FCGR2A* | (Nalls et al. 2019) |
| chr17:77929399-78929399 | *DNAH17* | (Nalls et al. 2019) |
| chr3:28164199-29164199 | *RBMS3* | (Nalls et al. 2019) |
| chr4:75689212-76689212 | *SCARB2* | (Nalls et al. 2019) |
| chr4:75776901-76776901 | *FAM47E* | (Nalls et al. 2019) |
| chr12:45525303-46525303 | *SLC38A1* | (Nalls et al. 2019) |
| chr10:119156173-120156173 | *BAG3* | (Nalls et al. 2019) |
| chr3:17820267-18820267 | *TBC1D5* | (Nalls et al. 2019) |
| chr6:132389222-133389222 | *HMGB1P13* | (Nalls et al. 2019) |
| chr2:17466582-18466582 | *KCNS3* | (Nalls et al. 2019) |
| chr1:154735843-155735843 | *GBA* | (Nalls et al. 2019) |
| chr12:39720632-40720632 | *LRRK2* | (Nalls et al. 2019) |
| chr7:66044864-67044864 | *AC006001.3* | (Nalls et al. 2019) |
| chr20:5525395-6525395 | *CRLS1* | (Nalls et al. 2019) |
| chr11:10037230-11037230 | *RNF141* | (Nalls et al. 2019) |

| | | |
|---|---|---|
| chr18:50657219-51657219 | *SMAD4* | (Nalls et al., 2019) |
| chr1:205254444-206254444 | *RAB29* | (Nalls et al., 2019) |
| chr17:43857262-44857262 | *FAM171A2* | (Nalls et al., 2019) |
| chr4:431588-1431588 | *GAK* | (Nalls et al., 2019) |
| chr10:15015407-16015407 | *ITGA8* | (Nalls et al., 2019) |
| chr6:29640906-30640906 | *TRIM40* | (Nalls et al., 2019) |
| chr13:48853596-49853596 | *CAB39L* | (Nalls et al., 2019) |
| chr14:87497920-88497920 | *GPR65* | (Nalls et al., 2019) |
| chr6:111422088-112422088 | *WISP3* | (Nalls et al., 2019) |
| chr1:154735878-155735878 | *GBA1* | (Rizig et al., 2023) |
| chr14:24713480-25147976 | *STXBP6* | (Loesch et al., 2021) |
| chr4:89695550-89866846 | *SNCA* | Known PD genes |
| chr6:161002496-163072863 | *PARK2* | Known PD genes |
| chr4:41253993-41271316 | *UCHL1* | Known PD genes |
| chr1:7955746-7991189 | *DJ-1* | Known PD genes |
| chr1:155264661-155274879 | *GBA* | Known PD genes |
| chr12:40188943-40405353 | *LRRK2* | Known PD genes |
| chr1:20628940-20656025 | *PINK1* | Known PD genes |
| chr1:16979465-17018421 | *ATP13A2* | Known PD genes |
| chr22:38093930-38199319 | *PLA2G6* | Known PD genes |
| chr22:32468692-32504859 | *FBXO7* | Known PD genes |
| chr3:184309279-184340574 | *EIF4G1* | Known PD genes |
| chr16:46652288-46696621 | *VPS35* | Known PD genes |
| chr1:226481196-226488644 | *ITPKB* | Known PD genes |
| chr2:101375382-101412082 | *IL1R2* | Known PD genes |
| chr2:164231020-164347533 | *SCN3A* | Known PD genes |
| chr3:18306448-18397281 | *SATB1* | Known PD genes |
| chr3:48636411-48648494 | *NCKIPSD* | Known PD genes |
| chr3:52164078-52180311 | *ALAS1* | Known PD genes |
| chr3:52187064-52192147 | *TLR9* | Known PD genes |
| chr3:52282303-52366475 | *DNAH1* | Known PD genes |
| chr3:52366997-52376334 | *BAP1* | Known PD genes |
| chr3:52376641-52389625 | *PHF7* | Known PD genes |
| chr3:52421102-52459052 | *NISCH* | Known PD genes |
| chr3:52461322-52490479 | *STAB1* | Known PD genes |
| chr3:52760782-52774993 | *ITIH3* | Known PD genes |
| chr3:52789919-52791298 | *ITIH4* | Known PD genes |
| chr4:111896953-112432557 | *ANK2* | Known PD genes |
| chr4:112529876-112840771 | *CAMK2D* | Known PD genes |

| | | |
|---|---|---|
| chr5:61455964-61548562 | *ELOVL7* | Known PD genes |
| chr6:27482964-27505339 | *ZNF184* | Known PD genes |
| chr8:11985015-12010720 | *CTSB* | Known PD genes |
| chr8:22694182-22718275 | *SORBS3* | Known PD genes |
| chr8:22721228-22736785 | *PDLIM2* | Known PD genes |
| chr8:22742134-22746637 | *C8orf58* | Known PD genes |
| chr8:22762905-22811607 | *BIN3* | Known PD genes |
| chr9:17579125-17797131 | *SH3GL2* | Known PD genes |
| chr10:15169644-15329060 | *FAM171A1* | Known PD genes |
| chr14:87466670-87526927 | *GALC* | Known PD genes |
| chr16:19056277-19068773 | *COQ7* | Known PD genes |
| chr16:52404096-52513096 | *TOX3* | Known PD genes |
| chr17:44381512-44445214 | *ATP6V0A1* | Known PD genes |
| chr17:44494947-44500427 | *PSMC3IP* | Known PD genes |
| chr17:44581937-44589638 | *TUBG2* | Known PD genes |
| chr3:87178135-87206349 | *CHMP2B* | Known PD genes |
| chr3:183298467-183300020 | *MCCC1* | Known PD genes |
| chr4:75326511-75385910 | *FAM47E* | Known PD genes |
| chr4:15680106-15688816 | *FAM200B* | Known PD genes |
| chr8:17169729-17265135 | *MICU3* | Known PD genes |
| chr10:117891859-117918308 | *BAG3* | Known PD genes |
| chr11:85179700-85179917 | *DLG2* | Known PD genes |
| chr11:134028609-134028686 | *MIR4697* | Known PD genes |
| chr12:39831272-39975479 | *LRRK2* | Known PD genes |
| chr12:122490673-122495494 | *OGFOD2* | Known PD genes |
| chr14:54375298-54436134 | *GCH1* | Known PD genes |
| chr14:67003549-67048754 | *TMEM229B* | Known PD genes |
| chr15:61560190-61768274 | *VPS13C* | Known PD genes |
| chr16:31063101-31072875 | *ZNF646* | Known PD genes |
| chr18:45687913-45697685 | *SYT4* | Known PD genes |
| chr19:2321518-2328621 | *LSM7* | Known PD genes |
| chr20:3209704-3224039 | *DDRGK1* | Known PD genes |
| chr17:47952550-48146446 | *KANSL1* | Known PD genes |
| chr17:47921884-47922328 | *STH* | Known PD genes |
| chr17:47767469-47769706 | *SPPL2C* | Known PD genes |
| chr17:47706954-47758460 | *CRHR1* | Known PD genes |
| chr17:47351987-47355550 | *ARHGAP27* | Known PD genes |
| chr16:31104433-31120072 | *KAT8* | Known PD genes |
| chr7:23206944-23235489 | *GPNMB* | Known PD genes |

| | | |
|---|---|---|
| chr4:15776655-15851607 | *CD38* | Known PD genes |
| chr2:167097510-167391631 | *STK39* | Known PD genes |
| chr7:23142208-23161390 | *NUPL2* | Known PD genes |
| chr7:23066165-23138295 | *KLHL7* | Known PD genes |
| chr6:32669654-32679285 | *HLA-DQA1* | Known PD genes |
| chr6:32584936-32592225 | *HLA-DRB6* | Known PD genes |
| chr4:965099-979768 | *DGKQ* | Known PD genes |
| chr2:134190255-134195627 | *CCNT2* | Known PD genes |
| chr4:938599-964868 | *TMEM175* | Known PD genes |
| chr2:133698188-133961429 | *TMEM163* | Known PD genes |
| chr1:232262219-232425812 | *SIPA1L2* | Known PD genes |
| chr1:205819965-205844048 | *SLC41A1* | Known PD genes |
| chr7:56101569-56106576 | *CHCHD2* | Known PD genes |
| chr20:3888839-3926958 | *PANK2* | Known PD genes |
| chr2:74361154-74392087 | *DCTN1* | Known PD genes |
| chr15:89316303-89334847 | *POLG1* | Known PD genes |
| chr1:151220451-153220451 | *HRNR* | Known PD genes |
| chr1:153925709-155928394 | *PMVK* | Known PD genes |
| chr1:154162560-156165245 | *KRTCAP2* | Known PD genes |
| chr1:154233158-156235843 | *GBAP1* | Known PD genes |
| chr1:154404265-156406949 | *ASH1L* | Known PD genes |
| chr1:155182384-157185070 | *SEMA4A* | Known PD genes |
| chr1:160499264-162499264 | *FCGR2A* | Known PD genes |
| chr1:170750628-172750629 | *VAMP4* | Known PD genes |
| chr1:206009146-206268644 | *NUCKS1* | Known PD genes |
| chr1:206009146-206268644 | *RAB29* | Known PD genes |
| chr1:225728376-227728377 | *ITPKB* | Known PD genes |
| chr1:231528865-233528865 | *SIPA1L2* | Known PD genes |
| chr2:16966581-18966570 | *KCNS3* | Known PD genes |
| chr9:40365644-40365693 | *KCNIP3* | Known PD genes |
| chr2:100780501-102780504 | *MAP4K4* | Known PD genes |
| chr2:133681218-135681219 | *TMEM163* | Known PD genes |
| chr2:167253884-169253884 | *STK39* | Known PD genes |
| chr3:17320267-19320267 | *SATB1* | Known PD genes |
| chr3:27664199-29664199 | *LINC00693* | Known PD genes |
| chr3:47707499-49711556 | *IP6K2* | Known PD genes |
| chr3:121478045-123478045 | *KPNA1* | Known PD genes |
| chr3:150391178-152391176 | *MED12L* | Known PD genes |
| chr3:160359842-162359842 | *SPTSSB* | Known PD genes |

| | | |
|---|---|---|
| chr3:182042285-184042285 | *MCCC1* | Known PD genes |
| chr4:849276-932373 | *GAK* | Known PD genes |
| chr4:932387-958656 | *TMEM175* | Known PD genes |
| chr4:14735724-16735725 | *BST1* | Known PD genes |
| chr4:16967188-18967188 | *LCORL* | Known PD genes |
| chr4:75185155-77189212 | *SCARB2* | Known PD genes |
| chr4:75222759-77226816 | *FAM47E* | Known PD genes |
| chr4:75272844-77276901 | *FAM47E-STBD1* | Known PD genes |
| chr4:88744890-90685975 | *SNCA* | Known PD genes |
| chr4:112447909-114447909 | *CAMK2D* | Known PD genes |
| chr4:168662006-170662006 | *CLCN3* | Known PD genes |
| chr5:59842133-61842132 | *ELOVL7* | Known PD genes |
| chr5:102030090-104030093 | *PAM* | Known PD genes |
| chr5:123774579-125774580 | *ZNF608* | Known PD genes |
| chr5:133863414-135863416 | *C5orf24* | Known PD genes |
| chr6:26737023-28771024 | *LOC100131289* | Known PD genes |
| chr6:29140906-31140906 | *TRIM40* | Known PD genes |
| chr6:30878457-32878457 | *SLC44A4* | Known PD genes |
| chr6:31610995-33610995 | *HLA-DRB5* | Known PD genes |
| chr6:70778059-72778039 | *RIMS1* | Known PD genes |
| chr6:110922088-112922089 | *FYN* | Known PD genes |
| chr6:131889221-133889223 | *RPS12* | Known PD genes |
| chr7:22260430-24260430 | *GPNMB* | Known PD genes |
| chr7:65544938-67544864 | *GS1-124K5.11* | Known PD genes |
| chr8:10854933-12854934 | *CTSB* | Known PD genes |
| chr8:15840084-17840084 | *FGF20* | Known PD genes |
| chr8:21668468-23668467 | *BIN3* | Known PD genes |
| chr8:128889663-130889663 | *FAM49B* | Known PD genes |
| chr9:16727067-18579692 | *SH3GL2* | Known PD genes |
| chr9:33046393-35046394 | *UBAP2* | Known PD genes |
| chr10:14515407-16515407 | *ITGA8* | Known PD genes |
| chr10:101255522-103255522 | *GBF1* | Known PD genes |
| chr10:118656173-120656173 | *BAG3* | Known PD genes |
| chr10:118776815-120776815 | *INPP5F* | Known PD genes |
| chr11:9537230-11537230 | *RNF141* | Known PD genes |
| chr11:82776235-84776234 | *DLG2* | Known PD genes |
| chr11:132917106-134917107 | *IGSF9B* | Known PD genes |
| chr12:31657825-33657824 | *FGD4* | Known PD genes |
| chr12:31826971-33826970 | *PKP2* | Known PD genes |

| | | |
|---|---|---|
| chr12:33792942-35252126 | *ALG10* | Known PD genes |
| chr12:39528770-41220632 | *LRRK2* | Known PD genes |
| chr12:40053867-41542837 | *CNTN1* | Known PD genes |
| chr12:40477002-42477002 | *PDZRN4* | Known PD genes |
| chr12:41141601-42901976 | *GXYLT1* | Known PD genes |
| chr12:45025303-47025303 | *SCAF11* | Known PD genes |
| chr12:121888692-123842051 | *HIP1R* | Known PD genes |
| chr12:132241105-132241213 | *FBRSL1* | Known PD genes |
| chr13:48353596-50353596 | *CAB39L* | Known PD genes |
| chr13:96212767-98212767 | *MBNL2* | Known PD genes |
| chr14:36520065-38520066 | *MIPOL1* | Known PD genes |
| chr14:53882151-55882151 | *GCH1* | Known PD genes |
| chr14:73906331-75906691 | *RPS6KL1* | Known PD genes |
| chr14:86997920-88997920 | *GALC* | Known PD genes |
| chr15:60705186-62705186 | *VPS13C* | Known PD genes |
| chr16:18183636-20266171 | *SYT17* | Known PD genes |
| chr16:27933075-29933075 | *CD19* | Known PD genes |
| chr16:29966478-31966478 | *SETD1A* | Known PD genes |
| chr16:49702745-51702745 | *NOD2* | Known PD genes |
| chr16:51602331-53602330 | *CASC16* | Known PD genes |
| chr16:51935514-53935514 | *CHD9* | Known PD genes |
| chr16:57553760-59553768 | *CNOT1* | Known PD genes |
| chr17:6452301-8452303 | *CHRNB1* | Known PD genes |
| chr17:41584761-43663645 | *RETREG3* | Known PD genes |
| chr17:43142320-45216970 | *UBTF* | Known PD genes |
| chr17:43357262-45357264 | *FAM171A2* | Known PD genes |
| chr17:44339905-46339907 | *MAP3K14* | Known PD genes |
| chr17:44720940-46666837 | *CRHR1* | Known PD genes |
| chr17:44855939-46855941 | *MAPT-AS1* | Known PD genes |
| chr17:45111699-47111701 | *KANSL1* | Known PD genes |
| chr17:45723580-47723580 | *NSF* | Known PD genes |
| chr17:45789439-47789439 | *WNT3* | Known PD genes |
| chr17:60840005-62840005 | *BRIP1* | Known PD genes |
| chr17:77429398-79429398 | *DNAH17* | Known PD genes |
| chr18:32724355-34724354 | *ASXL3* | Known PD genes |
| chr18:42093416-44093415 | *RIT2* | Known PD genes |
| chr18:50157219-52157219 | *MEX3C* | Known PD genes |
| chr19:1341048-3341049 | *SPPL2B* | Known PD genes |
| chr20:2184040-4184039 | *DDRGK1* | Known PD genes |

| | | |
|---|---|---|
| chr20:5025395-7025394 | *CRLS1* | Known PD genes |
| chr21:36480063-38480438 | *DYRK1A* | Known PD genes |
| chr16:74212955-75274831 | *FA2H* | Pallido-pyramidal syndrome (Simón-Sánchez et al., 2012) |
| chr21:43219094-44262307 | *PDXK* | Pallido-pyramidal syndrome (Simón-Sánchez et al., 2012) |
| chr15:44054818-45163688 | *SPG11* | Pallido-pyramidal syndrome (Simón-Sánchez et al., 2012) |
| chr6:146043833-147054953 | *RAB32* | Known PD gene (Hop et al., 2024) |
| chr20:613240-1689415 | *PSMF1* | Known PD gene (Magrinelli et al., 2024) |
| chr1:10507411-11507411 | *C1orf127* | Identified in SAPDSC GWAS |
| chr1:68665542-69665542 | *RP4-694A7.4* | Identified in SAPDSC GWAS |
| chr1:81620976-82620976 | *LPHN2* | Identified in SAPDSC GWAS |
| chr1:84833342-85833342 | *LPAR3* | Identified in SAPDSC GWAS |
| chr1:110596207-111596207 | *KCNA10* | Identified in SAPDSC GWAS |
| chr1:154751250-155751250 | *HCN3* | Identified in SAPDSC GWAS |
| chr1:159849417-160849417 | *NHLH1* | Identified in SAPDSC GWAS |
| chr1:206063867-207063867 | *SRGAP2* | Identified in SAPDSC GWAS |
| chr1:241075694-242075694 | *RP11-527D7.1* | Identified in SAPDSC GWAS |
| chr1:241173393-242173393 | *FH* | Identified in SAPDSC GWAS |
| chr1:241212134-242212134 | *KMO* | Identified in SAPDSC GWAS |
| chr2:7191515-8191515 | *RNU6ATAC37P* | Identified in SAPDSC GWAS |
| chr2:21595018-22595018 | *AC067959.1* | Identified in SAPDSC GWAS |
| chr2:24663010-25663010 | *DNAJC27* | Identified in SAPDSC GWAS |
| chr2:41420728-42420728 | *snoZ247* | Identified in SAPDSC GWAS |
| chr2:49620377-50620377 | *SNORA75* | Identified in SAPDSC GWAS |
| chr2:106181048-107181048 | *C2orf40* | Identified in SAPDSC GWAS |
| chr2:151750326-152750326 | *TNFAIP6* | Identified in SAPDSC GWAS |
| chr2:189415904-190415904 | *COL5A2* | Identified in SAPDSC GWAS |
| chr3:1276913-2276913 | *RPL23AP39* | Identified in SAPDSC GWAS |
| chr3:23912443-24912443 | *THRB* | Identified in SAPDSC GWAS |
| chr3:54787618-55787618 | *RP11-64C1.1* | Identified in SAPDSC GWAS |
| chr3:119213622-120213622 | *GSK3B* | Identified in SAPDSC GWAS |
| chr3:154388652-155388652 | *MME:MME-AS1* | Identified in SAPDSC GWAS |
| chr4:14404886-15404886 | *CPEB2-AS1* | Identified in SAPDSC GWAS |
| chr4:39994067-40994067 | *RBM47* | Identified in SAPDSC GWAS |
| chr4:55105943-56105943 | *KIT* | Identified in SAPDSC GWAS |
| chr4:110457107-111457107 | *ELOVL6* | Identified in SAPDSC GWAS |
| chr4:141224585-142224585 | *Y_RNA* | Identified in SAPDSC GWAS |

| | | |
|---|---|---|
| chr4:166297167-167297167 | *TLL1* | Identified in SAPDSC GWAS |
| chr4:169256160-170256160 | *PALLD* | Identified in SAPDSC GWAS |
| chr4:178872914-179872914 | *RNA5SP173* | Identified in SAPDSC GWAS |
| chr4:178891547-179891547 | *RNA5SP173* | Identified in SAPDSC GWAS |
| chr4:178902979-179902979 | *RNA5SP173* | Identified in SAPDSC GWAS |
| chr4:180557586-181557586 | *RP11-774G5.1* | Identified in SAPDSC GWAS |
| chr4:184969658-185969658 | *RP11-326I11.1* | Identified in SAPDSC GWAS |
| chr4:185088755-186088755 | *PRIMPOL* | Identified in SAPDSC GWAS |
| chr5:14428580-15428580 | *AC016575.1* | Identified in SAPDSC GWAS |
| chr5:68193328-69193328 | *RAD17* | Identified in SAPDSC GWAS |
| chr5:76754737-77754737 | *RP11-107N7.1* | Identified in SAPDSC GWAS |
| chr5:166374373-167374373 | *TENM2* | Identified in SAPDSC GWAS |
| chr5:173236333-174236333 | *CTC-430J12.2* | Identified in SAPDSC GWAS |
| chr6:9051830-10051830 | *OFCC1* | Identified in SAPDSC GWAS |
| chr6:52192440-53192440 | *GSTA5* | Identified in SAPDSC GWAS |
| chr6:53556273-54556273 | *MLIP* | Identified in SAPDSC GWAS |
| chr6:73313763-74313763 | *KCNQ5* | Identified in SAPDSC GWAS |
| chr6:137813241-138813241 | *RPSAP42* | Identified in SAPDSC GWAS |
| chr6:144695380-145695380 | *UTRN* | Identified in SAPDSC GWAS |
| chr6:151139564-152139564 | *AKAP12* | Identified in SAPDSC GWAS |
| chr7:39401576-40401576 | *AC072061.2* | Identified in SAPDSC GWAS |
| chr7:62190842-63190842 | *PHKG1P1* | Identified in SAPDSC GWAS |
| chr7:83027931-84027931 | *AC006322.1* | Identified in SAPDSC GWAS |
| chr7:88972272-89972272 | *STEAP2-AS1* | Identified in SAPDSC GWAS |
| chr7:94089716-95089716 | *PPP1R9A* | Identified in SAPDSC GWAS |
| chr7:98555898-99555898 | *PTCD1:ATP5J2-PTCD1:ATP5J2* | Identified in SAPDSC GWAS |
| chr7:101338531-102338531 | *CUX1* | Identified in SAPDSC GWAS |
| chr7:101883844-102883844 | *FAM185A* | Identified in SAPDSC GWAS |
| chr8:8164097-9164097 | *MFHAS1* | Identified in SAPDSC GWAS |
| chr8:11256600-12256600 | *RP11-589N15.1* | Identified in SAPDSC GWAS |
| chr8:12466965-13466965 | *DLC1* | Identified in SAPDSC GWAS |
| chr8:17045366-18045366 | *MTUS1* | Identified in SAPDSC GWAS |
| chr8:20857957-21857957 | *AC009695.1* | Identified in SAPDSC GWAS |
| chr8:37892182-38892182 | *C8orf86* | Identified in SAPDSC GWAS |
| chr8:53203840-54203840 | *RB1CC1* | Identified in SAPDSC GWAS |
| chr8:67246214-68246214 | *C8orf44-SGK3:SGK3* | Identified in SAPDSC GWAS |
| chr8:68461481-69461481 | *PREX2* | Identified in SAPDSC GWAS |
| chr8:72804334-73804334 | *RNA5SP271* | Identified in SAPDSC GWAS |
| chr8:93545866-94545866 | *TRIQK* | Identified in SAPDSC GWAS |

| | | |
|---|---|---|
| chr8:105823868-106823868 | *RP11-127H5.1* | Identified in SAPDSC GWAS |
| chr8:132223722-133223722 | *CTD-2008O4.1* | Identified in SAPDSC GWAS |
| chr8:132831630-133831630 | *KCNQ3* | Identified in SAPDSC GWAS |
| chr9:1483715-2483715 | *RP11-443B9.1* | Identified in SAPDSC GWAS |
| chr9:6933819-7933819 | *RPL4P5* | Identified in SAPDSC GWAS |
| chr9:6944090-7944090 | *RPL4P5* | Identified in SAPDSC GWAS |
| chr9:8466763-9466763 | *PTPRD* | Identified in SAPDSC GWAS |
| chr9:14006182-15006182 | *RP11-408A13.2* | Identified in SAPDSC GWAS |
| chr9:14586411-15586411 | *RP11-54D18.3* | Identified in SAPDSC GWAS |
| chr9:16260857-17260857 | *BNC2* | Identified in SAPDSC GWAS |
| chr9:17039496-18039496 | *CNTLN* | Identified in SAPDSC GWAS |
| chr10:60311409-61311409 | *RP11-561G1.3* | Identified in SAPDSC GWAS |
| chr10:110559517-111559517 | *RNU6-839P* | Identified in SAPDSC GWAS |
| chr10:111565249-112565249 | *SMNDC1* | Identified in SAPDSC GWAS |
| chr10:113993786-114993786 | *VTI1A* | Identified in SAPDSC GWAS |
| chr10:114643033-115643033 | *RNU7-165P* | Identified in SAPDSC GWAS |
| chr10:115233727-116233727 | *NHLRC2* | Identified in SAPDSC GWAS |
| chr10:126223133-127223133 | *CTBP2* | Identified in SAPDSC GWAS |
| chr11:6879100-7879100 | *SYT9* | Identified in SAPDSC GWAS |
| chr11:9892395-10892395 | *AMPD3* | Identified in SAPDSC GWAS |
| chr11:56479035-57479035 | *LRRC55* | Identified in SAPDSC GWAS |
| chr11:124167842-125167842 | *MSANTD2* | Identified in SAPDSC GWAS |
| chr12:9400366-10400366 | *CD69* | Identified in SAPDSC GWAS |
| chr12:98379174-99379174 | *RP11-181C3.1* | Identified in SAPDSC GWAS |
| chr12:121883451-122883451 | *WDR66* | Identified in SAPDSC GWAS |
| chr13:20538154-21538154 | *CRYL1* | Identified in SAPDSC GWAS |
| chr13:74227498-75227498 | *KLF12* | Identified in SAPDSC GWAS |
| chr13:81786469-82786469 | *PTMAP5* | Identified in SAPDSC GWAS |
| chr13:91903117-92903117 | *GPC5* | Identified in SAPDSC GWAS |
| chr13:95245248-96245248 | *ABCC4* | Identified in SAPDSC GWAS |
| chr13:98781073-99781073 | *CYCSP35* | Identified in SAPDSC GWAS |
| chr13:110355256-111355256 | *COL4A1* | Identified in SAPDSC GWAS |
| chr14:32297503-33297503 | *AKAP6* | Identified in SAPDSC GWAS |
| chr14:32492527-33492527 | *AKAP6* | Identified in SAPDSC GWAS |
| chr14:32553039-33553039 | *AKAP6* | Identified in SAPDSC GWAS |
| chr14:102040565-103040565 | *RN7SL472P* | Identified in SAPDSC GWAS |
| chr15:29092087-30092087 | *FAM189A1* | Identified in SAPDSC GWAS |
| chr15:31710943-32710943 | *SNORA18* | Identified in SAPDSC GWAS |
| chr15:35724678-36724678 | *MIR4510* | Identified in SAPDSC GWAS |

| | | |
|---|---|---|
| chr15:53193833-54193833 | *WDR72* | Identified in SAPDSC GWAS |
| chr15:54802249-55802249 | *RP11-548M13.1* | Identified in SAPDSC GWAS |
| chr15:77935064-78935064 | *IDH3A:RP11-285A1.1* | Identified in SAPDSC GWAS |
| chr15:91031872-92031872 | *PRC1* | Identified in SAPDSC GWAS |
| chr16:6446640-7446640 | *RBFOX1* | Identified in SAPDSC GWAS |
| chr16:9070529-10070529 | *RP11-418I22.2* | Identified in SAPDSC GWAS |
| chr16:9163057-10163057 | *RNA5SP403* | Identified in SAPDSC GWAS |
| chr16:23512413-24512413 | *PRKCB* | Identified in SAPDSC GWAS |
| chr16:23530819-24530819 | *PRKCB* | Identified in SAPDSC GWAS |
| chr16:78576084-79576084 | *WWOX* | Identified in SAPDSC GWAS |
| chr16:79025249-80025249 | *RP11-467I17.1* | Identified in SAPDSC GWAS |
| chr16:86432962-87432962 | *RP11-107C10.1* | Identified in SAPDSC GWAS |
| chr16:90064195-90338345 | *RPH3AL* | Identified in SAPDSC GWAS |
| chr17:1-725850 | *RPH3AL* | Identified in SAPDSC GWAS |
| chr17:3540741-4540741 | *ZZEF1* | Identified in SAPDSC GWAS |
| chr17:10152915-11152915 | *CTC-297N7.5:CTC-297N7.9* | Identified in SAPDSC GWAS |
| chr17:42517902-43517902 | *KIF18B* | Identified in SAPDSC GWAS |
| chr17:61893499-62893499 | *RPL31P57* | Identified in SAPDSC GWAS |
| chr18:56210939-57210939 | *OACYLP* | Identified in SAPDSC GWAS |
| chr18:56231725-57231725 | *OACYLP* | Identified in SAPDSC GWAS |
| chr19:8680458-9680458 | *OR1M4P* | Identified in SAPDSC GWAS |
| chr19:19986212-20986212 | *ZNF826P* | Identified in SAPDSC GWAS |
| chr19:31328834-32328834 | *TSHZ3* | Identified in SAPDSC GWAS |
| chr19:31735139-32735139 | *RNU6-967P* | Identified in SAPDSC GWAS |
| chr20:9816258-10816258 | *SNAP25-AS1* | Identified in SAPDSC GWAS |
| chr20:17716079-18716079 | *RP4-717M23.2* | Identified in SAPDSC GWAS |
| chr22:18785790-19785790 | *CLTCL1* | Identified in SAPDSC GWAS |
| chr22:31856243-32856243 | *YWHAH* | Identified in SAPDSC GWAS |
| Legend: chr, chromosome; PD, Parkinson's disease; SAPDSC; South African Parkinson's Disease Study Collection. Positions on GRCh38. | | |

**Supplementary Table 5:** Confusion matrix output to determine ancestral inference accuracy

| | AFR | EUR | MAL | NAMA | SAS |
|---|---|---|---|---|---|
| **AFR** | 33389 | 223 | 13 | 113 | 41 |
| **EUR** | 204 | 10112 | 18 | 36 | 107 |
| **MAL** | 43 | 126 | 1604 | 8 | 156 |
| **NAMA** | 258 | 121 | 61 | 3659 | 141 |
| **SAS** | 101 | 547 | 53 | 28 | 4796 |
| Legend: AFR, Africa; EUR, European; MAL, Malaysian; NAMA, Nama; SAS, South Asian | | | | | |

**Supplementary Table 6:** Top ten genome-wide association study hits gene implications according to Open Target Genetics

| rsID | Most severe consequence | Nearest gene(s) | Genes functionally implicated by this variant | Previous association with PD | Gene with PD association | Citation |
|---|---|---|---|---|---|---|
| rs190769295 | Intron variant | *ECRG4* | *NCK2, ECERG4, UXS1, RGPD3, CD8B2* | No | - | - |
| rs150739158 | Intron variant | *STXBP5-AS1* | *STXBP5, SAMD5* | No | - | - |
| rs562618836 | Intergenic variant | *RNU6-839P* | - | No | - | - |
| rs562970707 | Intergenic variant | *BTF3P15* | - | No | - | - |
| rs565473607 | Upstream gene variant | *SMNDC1* | - | No | - | - |
| rs6490582 | Intron variant | *CRYL1* | *CRYL1, GJB2, IFT88, IL17D, GJB6, EEF1AKMT1, GJA3, XPO4* | No | - | - |
| rs17098735 | Upstream gene variant | *AKAP6* | *AKAP6, ARHGAP5* | Yes | *AKAP6* | (Cappelletti et al. 2023; Phung et al. 2020) |
| rs8046041 | Regulatory region variant | *AC093519.2* | *FOXL1, MTHFSD, FOXC2, C16orf95, FOXF1, FBXO31, MAP1LC3B* | No | - | - |

| | | | | | | |
|---|---|---|---|---|---|---|
| rs75842871 | Intron variant | *KIF18B* | *KIF18B, NMT1, GFAP, CCDC103, FAM187A, C1QL1, EFTUD2, DCAKD, GJC1, HIGB1B, ADAM11, PLCD3, HEXIM2, ACBD4, DBF4B, CCDC43, MEIOC, SPATA32, FMNL1, MAP3K14, FZD2, GPATCH8, ARHGAP27, HEXIM1* | Yes | *ADAM11, PLCD3, EFTUD2, HEXIM2, MAP3K14, FZD2, ARHGAP27, GPATCH8, FMNL1, SPATA32, MEIOC, HIGD1B, GJC1, DBF4B, CCDC43, HEXIM1, ACBD4* | (Bandres-Ciga et al. 2019; Simón-Sánchez et al. 2009; Pickrell et al. 2016; International Parkinson Disease Genom...; Do et al. 2011; Nalls et al. 2014; Chang et al. 2017; Rodrigo and Nyholt 2021) |
| rs140681053 | Intron variant | *RPH3AL* | *RPH3AL, VPS53, DOC2B, TLCD3A, C17orf97, RFLNB, GEMIN4, GLOD4, SCGB1C2, MRM3, NXN, TIMM22* | No | - | - |
| Legend: PD, Parkinson's disease | | | | | | |

**Supplementary Table 7:** Variants identified as suggestively-significant in South African Parkinson's Disease Study Collection genome-wide association study and replicated in LARGE-PD cohort

| Variant | rsID | Nearest gene(s) | BETA (SAPDSC GWAS) | BETA (LARGE-PD) | P-value (SAPDSC GWAS) | P-value (LARGE-PD) | Effect direction |
|---|---|---|---|---|---|---|---|
| 7: 39861977: C/T | rs75284276 | *CDK13* | -1.4638 | -1.0012 | 1.53E-06 | 1.03E-02 | Aligned |
| 7: 63230464: A/C | rs56353559 | *ZNF722* | -1.4695 | -0.9729 | 2.08E-06 | 2.04E-02 | Aligned |
| 15: 78142722: C/G | rs7165154 | *CIB2* | -0.6206 | -0.3464 | 2.56E-06 | 2.80E-02 | Aligned |
| Legend: LARGE-PD, Latin American Research Consortium on the Genetics of Parkinson's Disease; SAPDSC, South African Parkinson's Disease Study Collection | | | | | | | |

**Supplementary Table 8:** Comparison of previously identified genome-wide association study hits in Parkinson's disease and corresponding p-values from present study

| Chromosome | Base position | Alleles | External beta | SAPDSC beta | SAPDSC P-value | Study | Directionality | GWAS Method |
|---|---|---|---|---|---|---|---|---|
| 16 | 30932927 | A/G | 0.0903 | 0.254648 | 4.10E-03 | Kim et al. 2023 | Aligned | LA-GWAS |
| 17 | 62048221 | C/T | 0.0800 | 0.291966 | 3.30E-02 | Kim et al. 2023 | Aligned | LA-GWAS |
| 17 | 82087210 | G/C | 0.0817 | -0.282987 | 1.10E-02 | Kim et al. 2023 | Opposite | LA-GWAS |
| 20 | 31833121 | T/C | -3.86E-09 | -0.239742 | 1.20E-02 | Kim et al. 2023 | Aligned | LA-GWAS |
| 4 | 15735725 | G/A | 0.104 | 0.19 | 4.04E-02 | Kim et al. 2023 | Aligned | LA-GWAS |
| 4 | 76289234 | T/A | -0.06 | -0.178212 | 4.67E-02 | Kim et al. 2023 | Aligned | LA-GWAS |
| 4 | 89710796 | C/A | 0.3439 | 0.642552 | 6.95E-06 | Kim et al. 2023 | Aligned | LA-GWAS |
| 8 | 11854934 | A/C | -0.093 | -0.245 | 1.16E-02 | Kim et al. 2023 | Aligned | LA-GWAS |
| 9 | 17726890 | C/T | 0.0913 | 0.241866 | 2.74E-02 | Kim et al. 2023 | Aligned | LA-GWAS |
| 1 | 205768611 | G/A | 0.1315 | 0.338055 | 6.85E-03 | Nalls et al. 2019 | Aligned | LA-GWAS |
| 12 | 122842051 | G/T | 0.1478 | 0.301446 | 2.51E-03 | Nalls et al. 2019 | Aligned | LA-GWAS |
| 16 | 30966478 | G/A | 0.091 | 0.264529 | 2.99E-03 | Nalls et al. 2019 | Aligned | LA-GWAS |
| 17 | 44216969 | A/C | -0.0631 | -2.01E-01 | 3.78E-02 | Nalls et al. 2019 | Aligned | LA-GWAS |
| 2 | 17966582 | A/T | -0.1104 | -0.562185 | 1.86E-03 | Nalls et al. 2019 | Aligned | LA-GWAS |
| 3 | 28664199 | T/C | -0.0658 | -2.49E-01 | 1.57E-02 | Nalls et al. 2019 | Aligned | LA-GWAS |
| 4 | 15735725 | G/A | 0.102 | 1.90E-01 | 4.04E-02 | Nalls et al. 2019 | Aligned | LA-GWAS |
| 4 | 76276901 | C/T | -0.0912 | -2.93E-01 | 1.52E-03 | Nalls et al. 2019 | Aligned | LA-GWAS |
| 4 | 89704960 | G/A | -0.2774 | -3.32E-01 | 1.62E-04 | Nalls et al. 2019 | Aligned | LA-GWAS |
| 8 | 11854934 | A/C | -0.088 | -2.45E-01 | 1.16E-02 | Nalls et al. 2019 | Aligned | LA-GWAS |
| 16 | 30932927 | A/G | 0.0903 | 2.40E-01 | 1.96E-02 | Kim et al. 2023 | Aligned | Conventional GWAS |
| 17 | 42570011 | A/G | -0.0594 | -2.29E-01 | 4.49E-02 | Kim et al. 2023 | Aligned | Conventional GWAS |

| | | | | | | | | |
|---|---|---|---|---|---|---|---|---|
| 4 | 76289234 | T/A | -0.06 | -2.12E-01 | 3.83E-02 | Kim et al. 2023 | Aligned | Conventional GWAS |
| 4 | 89710796 | C/A | 0.3439 | 6.26E-01 | 1.88E-04 | Kim et al. 2023 | Aligned | Conventional GWAS |
| 8 | 11854934 | A/C | -0.0883 | -2.65E-01 | 1.87E-02 | Kim et al. 2023 | Aligned | Conventional GWAS |
| 11 | 133917106 | G/T | 0.1073 | -2.84E-01 | 3.70E-02 | Nalls et al. 2019 | Opposite | Conventional GWAS |
| 16 | 30966478 | G/A | 0.0907 | 2.28E-01 | 2.63E-02 | Nalls et al. 2019 | Aligned | Conventional GWAS |
| 17 | 42588995 | T/C | -0.0642 | -2.51E-01 | 2.55E-02 | Nalls et al. 2019 | Aligned | Conventional GWAS |
| 2 | 17966582 | A/T | -0.1104 | -5.12E-01 | 1.10E-02 | Nalls et al. 2019 | Aligned | Conventional GWAS |
| 4 | 76276901 | C/T | -0.0912 | -2.94E-01 | 4.29E-03 | Nalls et al. 2019 | Aligned | Conventional GWAS |
| 4 | 89704960 | G/A | -0.2774 | -3.33E-01 | 7.57E-04 | Nalls et al. 2019 | Aligned | Conventional GWAS |
| 8 | 11854934 | A/C | -0.0930 | -2.65E-01 | 1.87E-02 | Nalls et al. 2019 | Aligned | Conventional GWAS |

Legend: The 31 SNPs that were previously identified in PD GWAS (Nalls et al., 2019 and Kim et al., 2023) with p-value < 0.05 in either the conventional GWAS or the LA-GWAS with ATT. ATT, Cochran-Armitage trend test; GWAS, genome-wide association study.

**Supplementary Table 9:** Local ancestry genome-wide association study genome-wide significant loci for the Tractor model

| Ancestry | Chromosome | Base position | Alleles | rsID | Nearest gene(s) | BETA | SE | P-value |
|---|---|---|---|---|---|---|---|---|
| ALL | 8 | 3698117 | C/T | rs2251850 | *RP11-875H7.2, WNT5A, LRTM1* | 1.480 | 0.805 | 4.14E-07 |
| | 8 | 3698448 | C/T | rs2449208 | *CSMD1* | 0.829 | 1.483 | 7.53E-06 |
| | 3 | 55401915 | A/G | rs1960758 | *CSMD1* | -2.239 | 1.277 | 9.70E-06 |
| AFR | 9 | 1314595 | C/A | rs4740539 | *RP11-459D20.1, LOC102723803, DMRT1, DMRT2, DMRT3* | 1.066 | 0.227 | 1.31E-06 |
| | 9 | 7419373 | A/G | rs2148353 | *RPL4P5, DMAC1* | 0.958 | 0.217 | 4.84E-06 |
| | 9 | 1220082 | A/G | rs58553697 | *RPS27AP14, DMRT1, DMRT2, DMRT3* | 0.929 | 0.215 | 7.53E-06 |

Legend: The results are sorted based on p-value threshold. AFR, African; ALL, all ancestries combined; SE, standard error

**Banner Authors: Global Parkinson's Genetics Program**

| Country | Name | Institution | Funders and Disclosures |
|---|---|---|---|
| **Algeria** | Yasser Mecheri | Centre Hospitalo-Universitaire Dr Benbadis Constantine | Nothing to declare |
| **Argentina** | Emilia M Gatto | Sanatorio de la Trinidad Mitre- INEBA | Nothing to declare |
| | Marcelo Kauffman | Hospital JM Ramos Mejia | Nothing to declare |
| | Federico Capparelli | Centro de Educación Médica e Investigaciones Clínicas Norberto Quirno | Nothing to declare |
| **Armenia** | Samson Khachatryan | Somnus Neurology Clinic | Nothing to declare |
| | Zaruhi Tavadyan | Somnus Neurology Clinic | Nothing to declare |
| | Mariam Isayan | Somnus Neurology Clinic | Nothing to declare |
| **Australia** | Claire E Shepherd | Neuroscience Research Australia | The Sydney Brain Bank is located at and supported by Neuroscience Research Australia |
| | Kishore Kumar | Garvan Institute of Medical Research and Concord Repatriation General Hospital | Paul Ainsworth Family Foundation |
| | Melina Ellis | Concord Hospital | Nothing to declare |
| | Miguel E. Rentería | QIMR Berghofer Medical Research Institute | The Australian Parkinson's Genetics Study is supported by the Shake It Up Australia Foundation and The Michael J. Fox Foundation for Parkinson's Research |

| | | | |
|---|---|---|---|
| | Sulev Koks | Murdoch University | Nothing to declare |
| | Simon Rowe | Neuroscience Research Australia | Nothing to declare |
| | Laura Rudaks | Neuroscience Research Australia | Nothing to declare |
| | Dennis Yeow | Neuroscience Research Australia | Nothing to declare |
| | Carolyn Sue | Neuroscience Research Australia | Nothing to declare |
| | Victor Flores Ocampo | QIMR Berghofer Medical Research Institute | Nothing to declare |
| **Austria** | Alexander Zimprich | Medical University Vienna Austria | Nothing to declare |
| **Azerbaijan** | Kanan Jafarov | Istanbul Klinik | Nothing to declare |
| **Belgium** | David Crosiers | University of Antwerp | Nothing to declare |
| **Brazil** | Artur F. Schumacher-Schuh | Universidade Federal do Rio Grande do Sul / Hospital de Clínicas de Porto Alegre | Nothing to declare |
| | Carlos Rieder | Federal University of Health Sciences of Porto Alegre | Nothing to declare |
| | Paula Saffie Awad | Universidade Federal do Rio Grande do Sul | Global Parkinson's Genetics Program |
| | Vitor Tumas | University of São Paulo | Nothing to declare |
| | Sarah Camargos | Universidade Federal de Minas Gerais | Nothing to declare |
| | Lucas Faria Costa | Universidade Federal de Minas Gerais | Nothing to declare |
| **Canada** | Edward A. Fon | Montreal Neurological Institute | Nothing to declare |
| | Oury Monchi | Institut universitaire de gériatrie de Montréal | CIHR, Brain Canada, Parkinson Canada |
| | Edward Fon | McGill University | Nothing to declare |
| | Robert Thibault | Aligning Science Across Parkinson's | Nothing to declare |

| | | | |
|---|---|---|---|
| | Ziv Gan-Or | McGill University | Nothing to declare |
| | Anthony Lang | University of Toronto | Nothing to declare |
| | Konstantin Senkevich | McGill University | Nothing to declare |
| **Chile** | Benjamin Pizarro Galleguillos | Universidad de Chile | Nothing to declare |
| | Marcelo Miranda | Fundación Diagnosis | Nothing to declare |
| | Maria Leonor Bustamante | Faculty of Medicine Universidad de Chile | Nothing to declare |
| | Patricio Olguin | Universidad de Chile | Nothing to declare |
| | Pedro Chana | CETRAM | Nothing to declare |
| | Elias Fernandez | Hospital Regional de Concepción | Nothing to declare |
| **China** | Beisha Tang | Central South University | Nothing to declare |
| | Huifang Shang | West China Hospital Sichuan University | Nothing to declare |
| | Jifeng Guo | Xiangya Hospital | Nothing to declare |
| | Piu Chan | Capital Medical University | Nothing to declare |
| | Wei Luo | Zhejiang University | Nothing to declare |
| | Zhenhua Liu | Xiangya Hospital, Central South University | Nothing to declare |
| **Colombia** | Gonzalo Arboleda | Universidad Nacional de Colombia | Nothing to declare |
| | Jorge Orozco | Fundación Valle del Lili | Nothing to declare |
| | Marlene Jimenez del Rio | University of Antioquia | Nothing to declare |
| **Costa Rica** | Alvaro Hernandez | University of Costa Rica | Nothing to declare |

| | | | |
|---|---|---|---|
| **Denmark** | Per Borghammer | Aarhus University | Nothing to declare |
| **Egypt** | Mohamed Salama | The American University in Cairo | The AUC/ ASRT/ DAAD |
| | Walaa A. Kamel | Beni-Suef University | Nothing to declare |
| **Ethiopia** | Yared Z. Zewde | Addis Ababa University | Nothing to declare |
| **France** | Alexis Brice | Paris Brain Institute | Nothing to declare |
| | Jean-Christophe Corvol | Sorbonne Université | Nothing to declare |
| | Mari Vidailhet | Salpêtriere Hospital (AP-HP), Sorbonne Université | Nothing to declare |
| **Georgia** | Mariam Kekenadze | Tbilisi State Medical University | Nothing to declare |
| | Irine Khatiashvili | S. Khechinashvili University Hospital | Nothing to declare |
| **Germany** | Ana Westenberger | University of Lübeck | Nothing to declare |
| | Anastasia Illarionova | Deutsches Zentrum für Neurodegenerative Erkrankungen | Nothing to declare |
| | Brit Mollenhauer | University Medical Center Göttingen | Nothing to declare |
| | Christine Klein | University of Lübeck | CK serves as a medical Advisor to Centogene on genetic testing reports in the field of movement disorders, except Parkinson's disease, and is a member of the Scientific Advisory Board of Retromer Therapeutics |
| | Eva-Juliane Vollstedt | University of Lübeck | Nothing to declare |
| | Franziska Hopfner | Department of Neurology, University Hospital, LMU Munich | Nothing to declare |

| | | |
|---|---|---|
| Günter Höglinger | Department of Neurology, University Hospital, LMU Munich | Nothing to declare |
| Harutyun Madoev | University of Lübeck | Nothing to declare |
| Joanne Trinh | University of Lübeck | Nothing to declare |
| Katja Lohmann | University of Lübeck | Nothing to declare |
| Lara M. Lange | University of Lübeck and University Medical Center Schleswig-Holstein | Nothing to declare |
| Manu Sharma | University of Tubingen | Dr. Sharma is further funded by the Michael J Fox Foundation, USA Genetic Diversity in PD Program: GAP-India Grant ID: 009411. |
| Sergiu Groppa | University of Mainz | Nothing to declare |
| Thomas Gasser | University of Tübingen | Nothing to declare |
| Zih-Hua Fang | The German Center for Neurodegenerative Diseases | Nothing to declare |
| Karl Heilbron | Charité - Universitätsmedizin Berlin | Former employee of 23andMe, Inc. Owns stock and/or stock options in 23andMe, Inc. |
| Theresa Luth | University of Luebeck | Nothing to declare |
| Wenhua Sun | University of Teubingen | Nothing to declare |
| Inke König | University of Lübeck | Nothing to declare |
| Daniela Berg | University Medical Center Schleswig-Holstein | Nothing to declare |
| Bernhard Haslinger | Technical University of Munich | Nothing to declare |
| Teresa Kleinz | University of Lübeck | Nothing to declare |

| | | | |
|---|---|---|---|
| | Norbert Brüggemann | University of Lübeck | Dr. Brüggemann received honaria from Abbvie, Esteve, Ipsen, Merz, Teva and Zambon |
| **Ghana** | Albert Akpalu | University of Ghana Medical School | Nothing to declare |
| **Greece** | Georgia Xiromerisiou | University of Thessaly | Nothing to declare |
| | Georgios Hadjigorgiou | University of Thessaly | Nothing to declare |
| | Ioannis Dagklis | Aristotle University of Thessaloniki | Nothing to declare |
| | Ioannis Tarnanas | Ionian University | Nothing to declare |
| | Leonidas Stefanis | Biomedical research Foundation of the Academy of Athens | PPMI2 (funded by MJFF), ALAMEDA (H2020 grant), funding by HFRI |
| | Maria Stamelou | Diagnostic and Therapeutic Centre HYGEIA Hospital | The non-profit organisation for scientific research in Parkinson’s disease and related disorders |
| | Efthymios Dadiotis | University of Thessaly | Nothing to declare |
| **Honduras** | Alex Medina | Hospital San Felipe | Nothing to declare |
| **Hong Kong** | Germaine Hiu-Fai Chan | Queen Elizabeth Hospital | Nothing to declare |
| | Nancy Ip | The Hong Kong University of Science and Technology | Nothing to declare |
| | Nelson Yuk-Fai Cheung | Queen Elizabeth Hospital | Nothing to declare |

| | | | |
|---|---|---|---|
| | Phillip Chan | The Hong Kong University of Science and Technology | Nothing to declare |
| | Xiaopu Zhou | The Hong Kong University of Science and Technology | Nothing to declare |
| **India** | Asha Kishore | Aster Medcity | Michael J. Fox Foundation |
| | Divya KP | Sree Chitra Tirunal Institute for Medical Sciences and Technology | Nothing to declare |
| | Pramod Pal | National Institute of Mental Health & Neurosciences | Nothing to declare |
| | Prashanth Lingappa Kukkle | Manipal Hospital | Nothing to declare |
| | Roopa Rajan | All India Institute of Medical Sciences | Nothing to declare |
| | Rupam Borgohain | Nizam's Institute Of Medical Sciences | Nothing to declare |
| **Iran** | Mehri Salari | Shahid Beheshti University of Medical Science | Nothing to declare |
| **Israel** | Tamara Shiner | Tel Aviv Sourasky Medical Center | Nothing to declare |
| | Avner Thaler | Tel Aviv Sourasky Medical Center | Nothing to declare |
| **Italy** | Andrea Quattrone | Magna Græcia University of Catanzaro | Nothing to declare |
| | Enza Maria Valente | University of Pavia | Nothing to declare |
| | Grazia Annesi | National Research Council | Nothing to declare |
| | Lucilla Parnetti | University of Perugia | Nothing to declare |
| | Micol Avenali | University of Pavia | Nothing to declare |
| | Monica Gagliardi | Magna Graecia University | Nothing to declare |
| | Tommaso Schirinzi | University of Rome Tor Vergata | Nothing to declare |

| | | | |
|---|---|---|---|
| | Caterina Galandra | IRCCS Mondino Foundation | Nothing to declare |
| | Anna De Rosa | University of Naples Federico II | Nothing to declare |
| **Japan** | Manabu Funayama | Juntendo University | Nothing to declare |
| | Nobutaka Hattori | Juntendo University faculty of medicine | Nothing to declare |
| | Tomotaka Shiraishi | Jikei University School of Medicine | Nothing to declare |
| **Kazakhstan** | Altynay Karimova | Institute of Neurology and Neurorehabilitation | Nothing to declare |
| | Gulnaz Kaishibayeva | Institute of Neurology and Neurorehabilitation | Nothing to declare |
| | Aigerim Utegenova | West Kazakhstan Marat Ospanov State Medical University | Nothing to declare |
| | Aigul Yemagambetova | West Kazakhstan Marat Ospanov State Medical University | Nothing to declare |
| | Vadim Akhmetzhanov | Medline medical center | Nothing to declare |
| | Seitzhan Aidarov | National Center for Neurosurgery | Nothing to declare |
| | Tautanova Raushan | Astana Medical University | Nothing to declare |
| | Bagyzhan Syzdykova Syzdykova | City multi-field hospital No. 1 | Nothing to declare |
| | Dinara Alzhanova | Astana Medical University | Nothing to declare |
| | Zhanybek Myrzayev | International University of Postgraduate Education | Nothing to declare |
| | Saltanat Abdraimova | South Kazakhstan Medical Academy | Nothing to declare |

| | | | |
|---|---|---|---|
| **Kyrgyzstan** | Cholpon Shambetova | Kyrgyz State Medical Academy | Nothing to declare |
| **Luxembourg** | Rejko Krüger | University of Luxembourg | Nothing to declare |
| | Patrick May | University of Luxembourg | Nothing to declare |
| **Malaysia** | Ai Huey Tan | University of Malaya | Nothing to declare |
| | Azlina Ahmad-Annuar | University of Malaya | Nothing to declare |
| | Mohamed Ibrahim Norlinah | Universiti Kebangsaan Malaysia | Nothing to declare |
| | Nor Azian Abdul Murad | UKM Medical Molecular Biology Institute | Nothing to declare |
| | Shahrul Azmin | Universiti Kebangsaan Malaysia Medical Centre | Nothing to declare |
| | Shen-Yang Lim | University of Malaya | Nothing to declare |
| | Wael Mohamed | International Islamic University | Nothing to declare |
| | Yi Wen Tay | University of Malaya | Nothing to declare |
| | Azalea Tenerife Pajo | University of Malaya | Nothing to declare |
| **Mexico** | Daniel Martinez-Ramirez | Tecnologico de Monterrey | Nothing to declare |
| | Mayela Rodriguez-Violante | Instituto Nacional de Neurologia y Neurocirugia | Nothing to declare |
| | Paula Reyes-Pérez | Universidad Nacional Autónoma de México | Global Parkinson's Genetics Program |
| | Alejandra Medina Rivera | Universidad Nacional Autónoma de México | Nothing to declare |

| | | | |
|---|---|---|---|
| | Nancy Monroy Jaramillo | Instituto Nacional de Neurología y Neurocirugía | Nothing to declare |
| **Mongolia** | Bayasgalan Tserensodnom | Mongolian National University of Medical Sciences | Nothing to declare |
| **Nepal** | Rajeev Ojha | Tribhuvan University | Nothing to declare |
| **Netherlands** | Wilma Van De Berg | Vanderbilt University Medical Center | Nothing to declare |
| | Bas Bleom | Radboud University | Nothing to declare |
| | Bart Van de Warrenburg | Radboud University Medical Center | Nothing to declare |
| | Lisette Charbonnier | Brain Research and Innovation Center | Nothing to declare |
| **New Zealand** | Tim J. Anderson | University of Otago | Health Research Council of New Zealand; Ministry of Business Innovation and Employment (New Zealand), Neurological Foundation of New Zealand |
| | Toni L. Pitcher | University of Otago | Funding - Health Research Council of New Zealand |
| **Nigeria** | Arinola Sanyaolu | University of Lagos | Nothing to declare |
| | Njideka Okubadejo | University of Lagos | Michael J Fox Foundation; Tertiary Education Trust Fund (TETFUND) National Research Fund |
| | Oluwadamilola Ojo | College of Medicine of the University of Lagos | Nothing to declare |
| | Simon Izuchukwu Ozomma | University of Calabar Teaching Hospital | Nothing to declare |

| | | | |
|---|---|---|---|
| | Kolawole Wahab | University of Ilorin | Nothing to declare |
| **Norway** | Lasse Pihlstrøm | Oslo University Hospital | Southeastern Regional Health Authority, Norway and Michael J. Fox Foundation |
| | Manuela Tan | Oslo University Hospital | Southeastern Norway Regional Health Authority Norway and Michael J. Fox Foundation |
| | Ingeborg Haugesag Lie | Oslo University Hospital | Nothing to declare |
| | Jodi Maple-Grødem | Stavanger University Hospital | Nothing to declare |
| **Pakistan** | Shoaib Ur-Rehman | University of Science and Technology Bannu | Nothing to declare |
| **Peru** | Mario Cornejo-Olivas | Universidad Cientifica del Sur | Michael J. Fox Foundation for Parkinson's Research and Aligning Science Across Parkinson's Initiative |
| **Philippines** | Maria Leila Doquenia | Metropolitan Medical Center | Nothing to declare |
| | Raymond Rosales | Metropolitan Medical Center | Nothing to declare |
| **Puerto Rico** | Angel Vinuela | University of Puerto Rico | Nothing to declare |
| **Russia** | Elena Iakovenko | Research Center of Neurology | Nothing to declare |
| | Anna Gareeva | Ufa Federal Research Center | Nothing to declare |
| | Gulnara Akhmadeeva | Ufa Scientific Center | Nothing to declare |

| | | | |
|---|---|---|---|
| | Irina Gilyazova | Russian Academy of Sciences / Bashkir State Medical University | Nothing to declare |
| **Saudi Arabia** | Bashayer Al Mubarak | King Faisal Specialist Hospital and Research Center | Nothing to declare |
| | Muhammad Umair | King Abdullah International Medical Research Center | Nothing to declare |
| **Singapore** | Eng-King Tan | National Neuroscience Institute | Singapore National Medical Research Council (MOH-OFLCG-000207) |
| | Jia Nee Foo | Nanyang Technological University | Singapore National Medical Research Council (MOH-000559) |
| | Elaine Chew | Nanyang Technological University | Nothing to declare |
| **Slovenia** | Vesna van Midden | Ljubljana University Medical Centre | Nothing to declare |
| **South Africa** | Ferzana Amod | University of KwaZulu-Natal | Nothing to declare |
| | Jonathan Carr | University of Stellenbosch | Nothing to declare |
| | Soraya Bardien | Stellenbosch University | National Research Foundation of South Africa[Grant Number 129249] |
| | Nikita Pillay | University of the Western Cape | Nothing to declare |
| | Kathryn Step | Stellenbosch University | Nothing to declare |
| **South Korea** | Beomseok Jeon | Seoul National University Hospital | Nothing to declare |
| | Yun Joong Kim | Yongin Severance Hospital | Nothing to declare |
| | Jung Hwan Shin | Seoul National University | Nothing to declare |
| | Joowon Jang | Seoul National University | Nothing to declare |

| | | | |
|---|---|---|---|
| **Spain** | Esther Cubo | Hospital Universitario Burgos | Nothing to declare |
| | Ignacio Alvarez | University Hospital Mutua Terrassa | Nothing to declare |
| | Janet Hoenicka | Institut de Recerca Sant Joan de Deu | Fondo de Investigación Sanitaria, Instituto Salud Carlos III, Grant PI019/00126 |
| | Katrin Beyer | Research Institute Germans Trias i Pujol | Nothing to declare |
| | Maria Teresa Periñan | Instituto de Biomedicina de Sevilla | Nothing to declare |
| | Pau Pastor | University Hospital Germans Trias i Pujol | Nothing to declare |
| | Ruben Fernandez-Santiago | Hospital Clínic de Barcelona | Nothing to declare |
| | Pilar Gómez Garre | Instituto de Biomedicina de Sevilla | Nothing to declare |
| | Pablo Mir | Instituto de Biomedicina de Sevilla | Nothing to declare |
| **Sudan** | Sarah El-Sadig | Faculty of medicine university of Khartoum | Nothing to declare |
| **Sweden** | Kajsa Brolin | Lund University | Nothing to declare |
| | Per Svenningsson | Karolinska Institute | Nothing to declare |
| | Maria Swanberg | Lund University | Nothing to declare |
| **Switzerland** | Christiane Zweier | Inselspital Bern, University of Bern | Nothing to declare |
| | Gerd Tinkhauser | University Hospital Bern | Nothing to declare |
| | Paul Krack | Inselspital Bern, University of Bern | Nothing to declare |
| **Taiwan** | Chin-Hsien Lin | National Taiwan University Hospital | Nothing to declare |
| | Hsiu-Chuan Wu | Chang Gung Memorial Hospital | Nothing to declare |

| | | | |
|---|---|---|---|
| | Pin-Jui Kung | National Taiwan University | Global Parkinson's Genetics Program |
| | Ruey-Meei Wu | National Taiwan University Hospital | Minister of Science and Technology, Taiwan Government; National Taiwan University; Parkinson foundation (USA); Michael J. Fox Foundation |
| | Yihru Wu | Chang Gung Memorial Hospital | Nothing to declare |
| **Tajikistan** | Ganieva Manizha | Avicenna Tajik State Medical University | Nothing to declare |
| **Tunisia** | Rim Amouri | National Institute Mongi Ben Hamida of Neurology | Nothing to declare |
| | Samia Ben Sassi | Mongi Ben Hmida National Institute of Neurology | Nothing to declare |
| **Turkey** | A. Nazlı Başak | Koç University | Kirac Foundation and Koc Univ. |
| | Gencer Genc | Şişli Etfal Training and Research Hospital | Nothing to declare |
| | Özgür Öztop Çakmak | Koç University | Nothing to declare |
| | Sibel Ertan | Koç University | Nothing to declare |
| **United Kingdom** | Alastair Noyce | Queen Mary University of London | Prof. Noyce reports grants from Parkinson's UK, Barts Charity, Cure Parkinson's, NIHR, Innovate UK, Virginia Keiley benefaction, Alchemab, Aligning |

| | | | |
|---|---|---|---|
| | | | Science Across Parkinson’s and Michael J Fox Foundation. Consultancy and personal fees from Astra Zeneca, AbbVie, Profile, Roche, Biogen, UCB, Bial, Charco Neurotech, uMedeor and Britannia. |
| | Alejandro Martínez-Carrasco | University College London | Global Parkinson's Genetics Program |
| | Anette Schrag | University College London | NIHR UCL/H Biomedical Research Centre |
| | Anthony Schapira | University College London | ASAP |
| | Camille Carroll | University of Plymouth | C Carroll receives salary from University of Plymouth, University Hospitals Plymouth NHS Trust and National Institute of Health Research; she has received advisory, consulting, and/or lecture fees from AbbVie, Bial, Lundbeck, Global Kinetics, Britannia and Medscape, and research funding from Parkinson's UK, Edmond J Safra Foundation, National Institute of Health Research and Cure Parkinson’s |

| | | | |
|---|---|---|---|
| | Donald Grosset | University of Glasgow | Tracking Parkinson's (J-1101) is funded by Parkinson's UK |
| | Eleanor J. Stafford | University College London | Nothing to declare |
| | Henry Houlden | University College London | Nothing to declare |
| | Huw R Morris | University College London | Dr Morris is employed by UCL. In the last 12 months he reports paid consultancy from Roche and Amylyx ; lecture fees/honoraria - BMJ, Kyowa Kirin, Movement Disorders Society. Research Grants from Parkinson’s UK, Cure Parkinson’s Trust, PSP Association, CBD Solutions, Drake Foundation, Medical Research Council, Michael J Fox Foundation. Dr Morris is a co-applicant on a patent application related to C9ORF72 - Method for diagnosing a neurodegenerative disease (PCT/GB2012/052140) |
| | John Hardy | University College London | Nothing to declare |
| | Kin Ying Mok | Univeristy College London | Nothing to declare |
| | Mie Rizig | University College London | Nothing to declare |

| | | | |
|---|---|---|---|
| | Nicholas Wood | University College London | ASAP- CRN |
| | Nigel Williams | Cardiff University | Parkinson's UK |
| | Olaitan Okunoye | University College London | Nothing to declare |
| | Rauan Kaiyrzhanov | University College London | Nothing to declare |
| | Rimona Weil | University College London | Nothing to declare |
| | Seth Love | University of Bristol | Medical Research Council (UK) MR/T018569/1 |
| | Simona Jasaityte | University College London | Nothing to declare |
| | Sumit Dey | Queen Mary University of London | The Michael J. Fox Foundation for Parkinson's Research |
| | Vida Obese | University College London | Nothing to declare |
| | Spencer Finch | Queen Mary University of London | Nothing to declare |
| | Valentina Escott-Price | Cardiff University | Nothing to declare |
| | Hamin Lee | St George's, University of London | Nothing to declare |
| | Roger Barker | University of Cambridge | Nothing to declare |
| | Mina Ryten | University College London | Nothing to declare |
| | Michele Hu | University of Oxford | Nothing to declare |
| | Laura Parkkinen | University of Oxford | Nothing to declare |
| | Kailash Bhatia | University College London | Nothing to declare |
| | Richard Walker | Northumbria Healthcare at NHS Foundation Trust | Nothing to declare |

| | | | |
|---|---|---|---|
| | Steve Gentleman | Imperial College London | Nothing to declare |
| | Thomas Warner | University College London | Nothing to declare |
| | David Burn | Newcastle University | Nothing to declare |
| | Christian Lambert | Imperial College London | Nothing to declare |
| | Caroline Williams-Gray | University of Cambridge | Nothing to declare |
| | Chris Morris | Newcastle University | Newcastle University, Alzheimer's Society, Alzheimer's Research UK, Medical Research Council |
| | Deborah Attuah | YLD | Nothing to declare |
| | Raquel Real | University College London | Nothing to declare |
| | Yen Tai | Imperial College London | Nothing to declare |
| **USA** | Alberto Espay | University of Cincinnati | Nothing to declare |
| | Alyssa O'Grady | The Michael J. Fox Foundation for Parkinson's Research | Nothing to declare |
| | Andrew B Singleton | National Institute on Aging | Michael J Fox Foundation for Parkinson's disease Research and Aligning Science Across Parkinson's Initiative |
| | Andrew K. Sobering | Augusta University / University of Georgia Medical Partnership | Nothing to declare |
| | Bernadette Siddiqi | The Michael J. Fox Foundation for Parkinson's Research | Nothing to declare |

| | | | |
|---|---|---|---|
| | Bradford Casey | The Michael J. Fox Foundation for Parkinson's Research | Nothing to declare |
| | Brian Fiske | The Michael J. Fox Foundation for Parkinson's Research | Nothing to declare |
| | Cabell Jonas | Mid-Atlantic Permanente Medical Group | Nothing to declare |
| | Carlos Cruchaga | Washington University | National Institutes of Health (R01AG044546 (CC), P01AG003991(CC, JCM), RF1AG053303 (CC), RF1AG058501 (CC), U01AG058922 (CC), RF1AG074007 (YJS)), the Chuck Zuckerberg Initiative (CZI), the Michael J. Fox Foundation (LI, CC), and the Department of Defense (LI- W81XWH2010849). The recruitment and clinical characterization of research participants at Washington University were supported by NIH P30AG066444 (JCM), P01AG03991(JCM), and P01AG026276(JCM). |
| | Caroline B. Pantazis | National Institutes of Health | Nothing to declare |
| | Charisse Comart | The Michael J. Fox Foundation for Parkinson's Research | Nothing to declare |
| | Claire Wegel | Indiana University | Nothing to declare |

| | | | |
|---|---|---|---|
| | Cornelis Blauwendraat | National Institutes of Health | Nothing to declare |
| | Dan Vitale | National Institutes of Health | Nothing to declare |
| | Deborah Hall | Rush University | Nothing to declare |
| | Dena Hernandez | National Institutes of Health | Nothing to declare |
| | Ejaz Shiamim | Kaiser Permanente | Nothing to declare |
| | Ekemini Riley | Coalition for Aligning Science | Nothing to declare |
| | Faraz Faghri | National Institutes of Health | F.F.'s participation in this research was supported in part by the Intramural Research Program of the NIH, National Institute on Aging (NIA), National Institutes of Health, Department of Health and Human Services; project number ZO1 AG000535, as well as the National Institute of Neurological Disorders and Stroke. F.F.'s participation in this project was part of a competitive contract awarded to Data Tecnica International LLC by the National Institutes of Health to support open science research. |

| | | | |
|---|---|---|---|
| | Geidy E. Serrano | Banner Sun Health Research Institute | Banner Sun Health Research Institute Brain and Body Donation Program of Sun City, Arizona for the provision of human biological materials. The Brain and Body Donation Program has been supported by the National Institute of Neurological Disorders and Stroke (U24 NS072026 National Brain and Tissue Resource for Parkinson's Disease and Related Disorders), the National Institute on Aging (P30 AG19610 and P30AG072980, Arizona Alzheimer's Disease Center), the Arizona Department of Health Services (contract 211002, Arizona Alzheimer's Research Center), the Arizona Biomedical Research Commission (contracts 4001, 0011, 05-901 and 1001 to the Arizona Parkinson's Disease Consortium) and the Michael J. |

| | | | |
|---|---|---|---|
| | | | Fox Foundation for Parkinson's Research |
| | Hampton Leonard | National Institute on Aging/National Institutes of Health | H.L.L is supported by a competitive contract awarded to Data Tecnica International LLC by the National Institutes of Health to support open science research |
| | Hirotaka Iwaki | Data Tecnica International | H.I is supported by a competitive contract awarded to Data Tecnica International LLC by the National Institutes of Health to support open science research |
| | Honglei Chen | Michigan State University | NIH/DoD/Parkinson Foundation/MSU Foundation/Gibby vs. Parky Foundation - No COI to disclose |
| | Ignacio F. Mata | Cleveland Clinic | Funding from MJFF and NIH |

| | | | |
|---|---|---|---|
| | Ignacio Juan Keller Sarmiento | Northwestern University | Nothing to declare |
| | Jared Williamson | Kaiser Permanente | Nothing to declare |
| | Jonggeol Jeff Kim | National Institutes of Health | Nothing to declare |
| | Joseph Jankovic | Baylor College of Medicine | Nothing to declare |
| | Joshua Shulman | Baylor College of Medicine / Texas Children's Hospital | Collection of samples and data at Baylor College of Medicine was supported by Huffington Foundation. |
| | Justin C. Solle | The Michael J. Fox Foundation for Parkinson's Research | Nothing to declare |
| | Kaileigh Murphy | The Michael J. Fox Foundation for Parkinson's Research | Nothing to declare |
| | Kamalini Ghosh Galvelis | Parkinson's Foundation | Nothing to declare |
| | Karen Nuytemans | University of Miami Miller School of Medicine | This work has been supported by the American Parkinson Disease Association and the Margaret Q. Landenberger Research Foundation. |
| | Karl Kieburtz | Beth Israel Deaconess Medical Center | Nothing to declare |
| | Kate Andersh | National Institute on Aging | Nothing to declare |
| | Katerina Markopoulou | North Shore University Health System | Nothing to declare |
| | Kenneth Marek | Institute for Neurodegenerative Disorders | Consultant for Michael J Fox Foundation, GE |

| | | | |
|---|---|---|---|
| | | | Healthcare, Roche, UCB, BIAL, Denali, Takeda, , Cerapsir, UCB, Biohaven, Neuron23, Aprinoia, Astellas, Calico, Genentech, Invicro |
| | Kristin S. Levine | Data Tecnica International | Nothing to declare |
| | Lana M. Chahine | University of Pittsburgh | Nothing to declare |
| | Laura Ibanez | Washington University | Nothing to declare |
| | Laurel Screven | National Institute on Aging | Nothing to declare |
| | Lauren Ruffrage | University of Alabama at Birmingham | Nothing to declare |
| | Lisa Shulman | University of Maryland | Nothing to declare |
| | Luca Marsili | University of Cincinnati | Nothing to declare |
| | Maggie Kuhl | The Michael J. Fox Foundation for Parkinson's Research | Nothing to declare |
| | Marissa Dean | University of Alabama at Birmingham | Dr. Dean is an investigator in studies funded by Abbvie, Inc., Hoffmann-La Roche, CHDI Foundation, Inc., Annexon, Inc., Retrophin, Inc, Neurocrine Biosciences, UniQure Biopharma B.V., Praxis Precision Medicines, Neuraly, Inc., Michael J. Fox Foundation for Parkinson’s Research, and US Army Medical Research |

| | | | |
|---|---|---|---|
| | | | and Material Command (grant#W81XWH-18-1-0508). In addition, Dr. Dean receives support through the Huntington's Disease Society of American Centers of Excellence program. |
| | Mary B Makarious | National Institutes of Health | Nothing to declare |
| | Matthew Farrer | University of Florida - Neurology | Nothing to declare |
| | Mathew Koretsky | National Institutes of Health | Nothing to declare |
| | Megan J. Puckelwartz | Northwestern University | Nothing to declare |
| | Miguel Inca-Martinez | Cleveland Clinic | Nothing to declare |
| | Mike A. Nalls | National Institutes of Health | M.A.N.'s participation in this project was part of a competitive contract awarded to Data Tecnica International LLC by the National Institutes of Health to support open science research. M.A.N. also currently serves as an advisor for Clover Therapeutics and Neuron23 Inc. |
| | Naomi Louie | The Michael J. Fox Foundation for Parkinson's Research | Nothing to declare |

| | | | |
|---|---|---|---|
| | Niccolò Emanuele Mencacci | Northwestern University | Nothing to declare |
| | Roger Albin | Universit of Michigan | P50NS123067; Parkinson's Foundation |
| | Roy Alcalay | Columbia University | Dr. Alcalay is funded by the Micharl J. Fox Foundation and the Parkinson's Foundation. He received consultation fees from Avrobio, Caraway, GSK, Merck, Sanofi, Ono Therapeutics and Takeda |
| | Ruth Walker | James J. Peters Veterans Affairs Medical Center | Research funding from the Department of Veterans Affairs (CSRD Merit Award CX002342), consulted for Teladoc, Inc. and received an honorarium from New York University (NYU) Medical Center. |
| | Sara Bandres-Ciga | National Institutes of Health | Nothing to declare |
| | Sohini Chowdhury | The Michael J. Fox Foundation for Parkinson's Research | Nothing to declare |
| | Sonya Dumanis | Aligning Science Across Parkinson's | Nothing to declare |
| | Steven Lubbe | Northwestern University | Nothing to declare |
| | Tao Xie | University of Chicago | Nothing to declare |

| | | | |
|---|---|---|---|
| | Tatiana Foroud | Indiana University School of Medicine | The Michael J. Fox Foundation for Parkinson's Research |
| | Thomas Beach | Sun Health Research Institution | Banner Sun Health Research Institute Brain and Body Donation Program of Sun City, Arizona for the provision of human biological materials. The Brain and Body Donation Program has been supported by the National Institute of Neurological Disorders and Stroke (U24NS072026 National Brain and Tissue Resource for Parkinson’s Disease and Related Disorders), the National Institute on Aging (P30 AG19610 and P30AG072980, Arizona Alzheimer’s Disease Center), the Arizona Department of Health Services (contract 211002, Arizona Alzheimer’s Research Center), the Arizona Biomedical Research Commission (contracts 4001, 0011, 05-901 and 1001 to the Arizona Parkinson's Disease Consortium) and the |

| | | | |
|---|---|---|---|
| | | | Michael J. Fox Foundation for Parkinson’s Research |
| | Todd Sherer | The Michael J Fox Foundation for Parkinson's Research | Nothing to declare |
| | Yeajin Song | National Institutes of Health | Nothing to declare |
| | Dana Lewis | Aligning Science Across Parkinson's | Nothing to declare |
| | Shreya Menon | Gladstone Institutes | Nothing to declare |
| | Melissa Nirenberg | Icahn School of Medicine at Mount Sinai | Nothing to declare |
| | Spencer Grant | National Institutes of Health | Nothing to declare |
| | Shannon Ballard | National Institutes of Health | Nothing to declare |
| | Chad Shaw | Baylor College of Medicine | Nothing to declare |
| | Sidra Aslam | Banner Health | Nothing to declare |
| | Geidy Serrano | Banner Sun Health Research Institute | Nothing to declare |
| | Devin Sharp | Aligning Science Across Parkinson's | Nothing to declare |
| | Kensuke Daida | National Institute of Aging | Nothing to declare |
| | Rachel Saunders-Pullman | Icahn School of Medicine at Mount Sinai | Nothing to declare |
| | Michiko Kimura Bruno | The Queen's Medical Center | Nothing to declare |
| | Matt Farrer | University of Florida College of Medicine | Nothing to declare |

| | | | |
|---|---|---|---|
| | Haydeh Payami | The University of Alabama at Birmingham Heersink School of Medicine | Nothing to declare |
| | Ryan Pfingst | The Michael J Fox Foundation | Nothing to declare |
| | James B Leverenz | Cleveland Clinic | Nothing to declare |
| | Elizabeth Disbrow | LSU Health Shreveport | Nothing to declare |
| | Debi Brooks | The Michael J Fox Foundation | Nothing to declare |
| | Randy Schekman | University of California, Berkeley | Nothing to declare |
| | Un Kang | NYU Grossman School of Medicine | Nothing to declare |
| | Zbigniew K. Wszolek | Mayo Clinic College of Medicine | Nothing to declare |
| | Cyrus Zabetian | VA Puget Sound Health Care System | Nothing to declare |
| | Zach Chaney | The Michael J Fox Foundation | Nothing to declare |
| | Mark Cookson | National Institute of Health | Nothing to declare |
| | Christine Swanson-Fischer | National Institute of Health | Nothing to declare |
| | Conor Hennessey | The Michael J Fox Foundation | Nothing to declare |
| | Cassandra Barrett | The Michael J Fox Foundation | Nothing to declare |
| | Beate Ritz | University of California, Los Angeles | Nothing to declare |
| | Bradley Boeve | Mayo Clinic | Nothing to declare |
| | Ashley Rawls | University of Florida College of Medicine | Nothing to declare |
| **Vietnam** | Duan Nguyen | Hue University | Nothing to declare |
| | Toan Nguyen | Hue University | Nothing to declare |

| | | | |
|---|---|---|---|
| **Zambia** | Masharip Atadzhanov | University of Zambia | Nothing to declare |
